# Immunological imprinting shapes the specificity of human antibody responses against SARS-CoV-2 variants

**DOI:** 10.1101/2024.01.08.24301002

**Authors:** Timothy S. Johnston, Shuk Hang Li, Mark M. Painter, Reilly K. Atkinson, Naomi R. Douek, David B. Reeg, Daniel C. Douek, E. John Wherry, Scott E. Hensley

## Abstract

The spike glycoprotein of severe acute respiratory syndrome coronavirus-2 (SARS-CoV-2) continues to accumulate substitutions, leading to breakthrough infections of vaccinated individuals and prompting the development of updated booster vaccines. Here, we determined the specificity and functionality of antibody and B cell responses following exposure to BA.5 and XBB variants in individuals who received ancestral SARS-CoV-2 mRNA vaccines. BA.5 exposures elicited antibody responses that primarily targeted epitopes conserved between the BA.5 and ancestral spike, with poor reactivity to the XBB.1.5 variant. XBB exposures also elicited antibody responses that targeted epitopes conserved between the XBB.1.5 and ancestral spike. However, unlike BA.5, a single XBB exposure elicited low levels of XBB.1.5-specific antibodies and B cells in some individuals. Pre-existing cross-reactive B cells and antibodies were correlated with stronger overall responses to XBB but weaker XBB-specific responses, suggesting that baseline immunity influences the activation of variant-specific SARS-CoV-2 responses.

**Highlights:**

- Variant breakthrough infections boost ancestral cross-reactive antibodies and B cells
- First and second BA.5 exposures fail to elicit variant-specific antibodies and B cells
- XBB infections and monovalent vaccinations elicit XBB.1.5-specific responses in some individuals
- XBB.1.5-specific responses correlate with low levels of pre-existing humoral immunity

## Introduction

SARS-CoV-2 first emerged in 2019, prompting the rapid development of mRNA-LNP vaccines that elicit potent neutralizing antibodies targeting the viral spike glycoprotein (Corbett et al., Hseih et al., Jackson et al., 2020). The virus has since acquired many spike substitutions that prevent the binding of antibodies elicited by vaccinations and infections. Notably, the BA.1 Omicron variant that began spreading widely in late 2020 possessed ∼32 spike amino acid substitutions compared to the ancestral SARS-CoV-2 strain (Mannar et al., 2022), leading to an increase in ‘breakthrough’ infections of vaccinated individuals (Accorsi et al., Altarawneh et al., Cao et al., Kuhlmann et al., 2022; Levin et al., 2021; Pulliam et al., 2022).

Omicron breakthrough infections induce efficient anamnestic immune responses that recruit memory T and B cells cross-reactive to the ancestral strain (Addetia et al., 2023; Kared et al., Koutsakos et al., 2022; Painter et al., 2023; Park et al., Wang et al., 2022; Weber et al., Yisimayi et al., 2023). Many unanswered questions remain on how initial SARS-CoV-2 encounters affect the specificity of antibodies elicited against variant viral strains. For example, bivalent boosters containing BA.5 spike elicit B cell and antibody responses cross-reactive to the ancestral SARS-CoV-2 strain in individuals who received ancestral mRNA-LNP vaccines (Addetia et al., 2023), but this ‘immunological imprinting’ effect may be less pronounced in individuals who initially received inactivated vaccines (Yisimayi et al., 2023). It remains unclear if repeat exposures with SARS-CoV-2 variants can overcome memory B cell biases established by initial SARS-CoV-2 encounters (Addetia et al., Alsoussi et al., Schiepers et al., Yisimayi et al., 2023). Further, it is unknown if variants with larger antigenic distances are better able to stimulate *de novo* responses while recalling fewer memory B cells in individuals who received ancestral mRNA-LNP vaccines.

Here, we elucidated the specificity of antibody and B cell responses elicited by BA.5 and XBB exposures in individuals previously vaccinated with mRNA-LNPs expressing the ancestral SARS-CoV-2 spike. We compared immune responses in individuals with different amounts of cross-reactive B cells and antibodies at time of variant exposure. We used antigen-specific flow cytometric analyses to interrogate spike-specific B cell responses and performed enzyme-linked immunosorbent assays (ELISAs), neutralization assays, and absorption assays to characterize SARS-CoV-2 reactive antibodies.

## Results

### BA.5 breakthrough infections elicit antibodies that cross-react to ancestral SARS-CoV-2

To better understand the specificity and functionality of antibodies elicited by breakthrough infections, we characterized antibodies in sera collected from individuals (n=8) who received 3 doses of mRNA-LNP vaccines expressing the ancestral spike and were subsequently infected with a BA.5 Omicron variant in 2022 (breakthrough infections occurred on average 291 days since last vaccination; Painter et al., 2023) (**Figure 1A**). Sera were collected at baseline ∼0-5 days (T1) and ∼45 days (T2) after BA.5 breakthrough infection. We first quantified serum antibodies reactive to full length spike proteins from the ancestral virus, the breakthrough variant (BA.5), and a variant from 2023 that did not yet exist at the time of sample collection (XBB.1.5). Antibodies reactive to all 3 spike proteins increased >2 fold after BA.5 breakthrough infection (**Figure 1B**). Similar increases were observed when we measured antibodies reactive to the receptor binding domain (RBD) of the spike protein from the ancestral, BA.5, and XBB.1.5 viruses, with the largest fold increase to BA.5 RBD (**Figure 1C**). Prior to breakthrough infection, most participants had high neutralizing antibodies against ancestral SARS-CoV-2, but low or undetectable levels of neutralizing antibodies against BA.5 and XBB.1.5 (**Figure 1D**). gNeutralizing antibodies against ancestral SARS-CoV-2 and the BA.5 variant were boosted upon BA.5 variant breakthrough infection in most participants, whereas neutralizing antibodies against XBB.1.5 were minimally boosted and remained low after infection (**Figure 1D**). This led to an observed increase in antibody neutralization potency (the neutralizing antibody titer divided by total spike targeting antibody titer) for ancestral SARS-CoV-2 and BA.5, but not XBB.1.5 (**Figure 1E**). Thus, BA.5 breakthrough infections elicited neutralizing antibodies against ancestral SARS-CoV-2 and BA.5, but these antibodies poorly neutralized the XBB.1.5 variant that did not yet exist at the time of infection.

**Figure 1.**
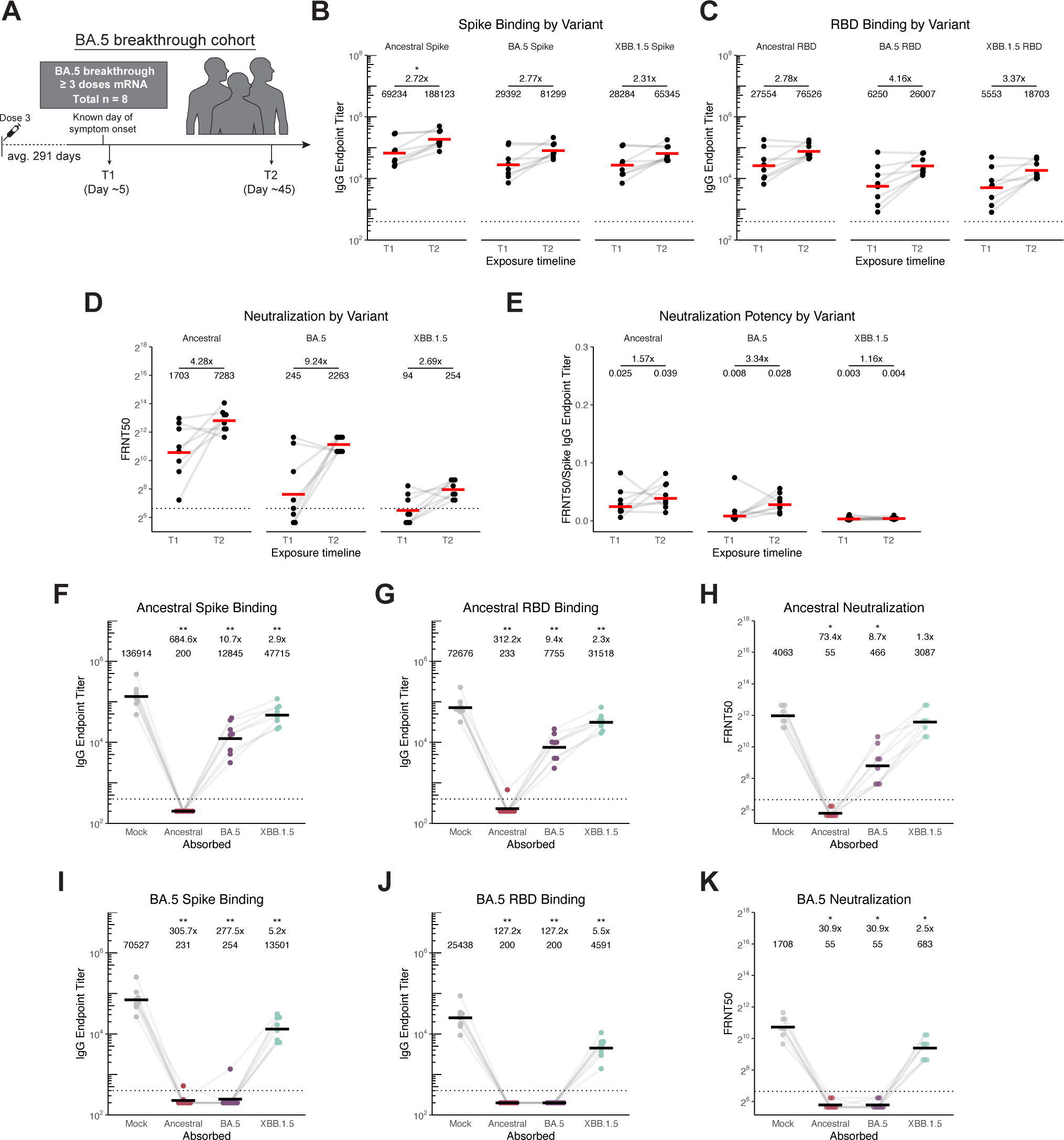
BA.5 breakthrough infection elicits cross-reactive antibodies evaded by XBB.1.5. A) Schematic of participants in this study who were vaccinated 3x with the SARS-CoV-2 ancestral mRNA-LNP vaccine who then had a BA.5 breakthrough infection. B)-C) Antigen-specific IgG ELISAs were performed using sera from timepoints indicated in panel A) against ancestral, BA.5, and XBB.1.5 full-length spike (B) and RBD (C) proteins. Endpoint titers are reported as reciprocal serum dilutions. D) SARS-CoV-2 pseudotype neutralization assays were performed using sera obtained at timepoints indicated in panel A) against ancestral SARS-CoV-2, BA.5, and XBB.1.5 pseudoviruses. Values reported are focus reduction neutralization test (FRNT) 50, or reciprocal serum dilution at which <50% viral input foci are observed. E) Neutralization potency was calculated by dividing FRNT50 values by spike IgG titer. F-G) Antigen-specific IgG ELISAs were performed using T2 absorbed sera and ELISA plates coated with the ancestral spike (F) or ancestral RBD (G). H) SARS-CoV-2 pseudotype neutralization assays were performed using T2 absorbed serum and ancestral SARS-CoV-2 pseudovirus. I-J) Antigen-specific IgG ELISAs were performed using T2 absorbed and ELISA plates coated with BA.5 spike (I) and BA.5 RBD (J). K) Neutralization assays were performed using T2 absorbed serum and BA.5 pseudotyped virus. For all, individual points are average of n = 2 technical replicates. Red/black bars indicate geometric mean. Wilcoxon signed-rank test with benjamini-hochberg correction for multiple testing. All comparisons to timepoint 1 or mock absorption. * p<0.05, ** p<0.01.

Given that we observed simultaneous increases in neutralizing antibodies reactive to the ancestral SARS-CoV-2 virus and BA.5 variant, we hypothesized that BA.5 breakthrough infections elicit neutralizing antibodies that recognize epitopes conserved between these viruses. To address this, we performed ELISAs and neutralization assays with sera that were previously absorbed with carboxyl-magnetic beads coupled with different spike proteins. In these assays (Anderson et al., 2022; Arevalo et al., 2020), antibodies that bind to the spike-coupled beads are removed and unbound antibodies are then assessed for reactivity against different antigens (**Figure S1A**). As expected, beads coated with the ancestral SARS-CoV-2 spike were able to remove >99% of antibodies reactive to the ancestral SARS-CoV-2 full length spike and RBD, while beads coated with either BA.5 or XBB.1.5 spikes reduced but did not entirely remove these antibodies (**Figure 1F-G**). Consistent with this observation, sera absorbed with beads coated with the ancestral SARS-CoV-2 spike could no longer neutralize ancestral SARS-CoV-2, while sera absorbed with beads coated with either the BA.5 or XBB.1.5 spike still contained antibodies capable of neutralizing the ancestral strain (**Figure 1H**). Conversely, beads coated with either the ancestral SARS-CoV-2 spike or BA.5 spike efficiently removed antibodies that bound (**Figure 1I-J**) and neutralized (**Figure 1K**) BA.5 virus, suggesting that nearly all BA.5-reactive antibodies elicited by BA.5 breakthrough infections cross-react to the ancestral SARS-CoV-2 spike. Throughout these studies, it became apparent that some of these ancestral/BA.5 cross-reactive antibodies recognize epitopes that are altered in XBB.1.5, since serum pre-incubation with beads coated with the XBB.1.5 spike did not eliminate binding (∼20% remaining, **Figure 1I-J**) or neutralization (∼40% remaining, **Figure 1K**) of BA.5. These data, combined with overall low XBB.1.5 titers (**Figure 1D, S1B-D**) suggest that BA.5 breakthrough infections elicit antibodies that cross-react with the ancestral SARS-CoV-2 and BA.5 spikes, and that substitutions in the XBB.1.5 variant prevent binding of a fraction of these antibodies, especially neutralizing antibodies.

### A second BA.5 exposure elicits antibodies that cross-react to ancestral SARS-CoV-2

We next analyzed sera from an additional 9 individuals who received 3 doses of an mRNA-LNP vaccine expressing the ancestral SARS-CoV-2 spike who were later exposed twice to BA.5 through sequential infections and vaccinations (**Figure 2A**). These individuals received a dose of bivalent ancestral/BA.5 spike mRNA-LNP vaccine either before (n=4) or after (n=5) experiencing a BA.5 breakthrough infection. Sera were collected early (∼0-5 days, T1) and ∼45 days (T2) after the second BA.5 exposure, which occurred on average 76 days (27-156) after the first BA.5 exposure. Antibodies reactive to the ancestral, BA.5, and XBB.1.5 spike proteins were high prior to the second BA.5 exposure and were minimally boosted (**Figure 2B-C**). Most individuals had high baseline neutralizing antibodies against both ancestral SARS-CoV-2 and BA.5 and these antibodies were minimally boosted upon second BA.5 exposure (**Figure 2D**). Neutralizing antibody titers against the XBB.1.5 variant were low in most individuals both before and after the second BA.5 exposure (**Figure 2D**). We observed a modest decrease in serum neutralization potency for all variants tested after secondary BA.5 exposure (**Figure 2E**). Antibody responses were similar regardless of whether the first or second BA.5 exposure was via infection or vaccination (**Figure S2A-C**).

**Figure 2.**
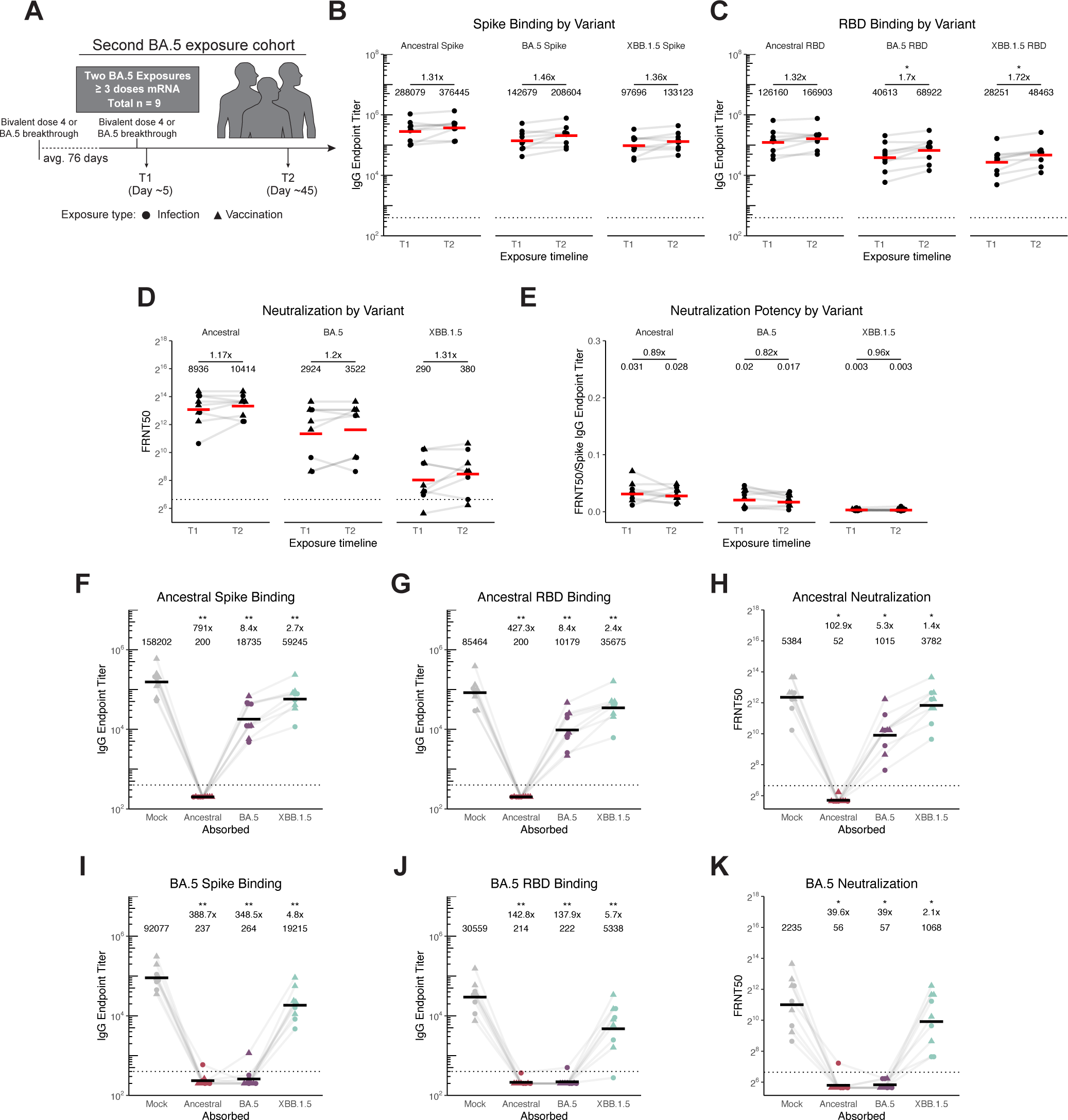
Secondary BA.5 exposure elicits cross-reactive antibodies that bind weakly to XBB.1.5. A) Schematic of participants in this study who were vaccinated 3x with the SARS-CoV-2 ancestral mRNA-LNP vaccine who then had two BA.5 exposures. B)-C) Antigen-specific IgG ELISAs were performed using sera from timepoints indicated in panel A) against ancestral, BA.5, and XBB.1.5 full-length spike (B) and RBD (C). Endpoint titers are reported as reciprocal dilutions. D) SARS-CoV-2 pseudotype neutralization assays were performed using sera obtained at timepoints indicated in panel A) against ancestral SARS-CoV-2, BA.5, and XBB.1.5 pseudoviruses. Values reported are focus reduction neutralization test (FRNT) 50, or reciprocal serum dilution at which <50% viral input foci are observed. E) Neutralization potency was calculated by dividing FRNT50 values by spike IgG titer. F)-G) Antigen-specific IgG ELISAs were performed using T2 absorbed sera and ELISA plates coated with the ancestral spike (F) or ancestral RBD (G). H) SARS-CoV-2 pseudotype neutralization assays were performed using T2 absorbed serum and ancestral SARS-CoV-2 pseudovirus. I-J) Antigen-specific IgG ELISAs were performed using T2 absorbed sera and ELISA plates coated with BA.5 spike (I) and BA.5 RBD (J). K) Neutralization assays were performed using T2 absorbed serum and BA.5 pseudotyped virus. For all, Individual points are average of n = 2 technical replicates. Red/black bars indicate geometric mean. Wilcoxon signed-rank test with benjamini-hochberg correction for multiple testing. All comparisons to timepoint 1 or mock absorption. * p<0.05, ** p<0.01.

We performed additional absorption assays and found that, similar to antibodies elicited by single BA.5 breakthrough infections, BA.5-reactive antibodies from individuals exposed twice with BA.5 bound to the ancestral SARS-CoV-2 spike, but only partially recognized the XBB.1.5 spike (**Figure 2F-K, S2D-F**). Thus, antibodies elicited by a second BA.5 spike exposure did not target BA.5-specific epitopes, but instead recognized epitopes conserved in the ancestral spike that became mutated in the XBB.1.5 variant.

### Cross-reactive B cell responses dominate first and second BA.5 exposures in individuals previously vaccinated with mRNA-LNP vaccines expressing the ancestral spike

We hypothesized that cross-reactive antibodies in our study were likely produced by B cells generated after the initial exposures to the ancestral spike and subsequently recalled in response to first and second BA.5 exposures. To measure levels of cross-reactive B cells elicited by first and second BA.5 exposures, we performed a flow cytometric B-cell antigen probe assay using PBMCs collected from participants at timepoints corresponding to our serological assays (**Fig. 3A, S3**). We quantified B cells that recognized the RBD of ancestral, BA.5, and XBB.1.5 spike proteins. Following first BA.5 exposures, the proportion of ancestral RBD-binding B cells that also bound BA.5 RBD increased in most individuals, and these responses did not greatly increase following a second BA.5 exposure (**Fig. 3B**). In further accordance with our serological data, XBB.1.5 cross-reactivity was only observed in a fraction of BA.5-binding B cells at all timepoints, and XBB.1.5 cross-reactivity did not increase following BA.5 infection (**Fig. 3C-D**). We did not observe robust variant-specific B cell responses, as extremely low frequencies of B cells bound BA.5 or XBB.1.5 RBD without binding ancestral RBD (BA.5-specific and XBB.1.5-specific, respectively), and the frequency of BA.5-only and XBB.1.5-only B cells did not increase following BA.5 exposure (**Fig. 3E-F**). Together, these data demonstrate that ancestral cross-reactive B cells are stimulated upon BA.5 exposure, leading to a heavily cross-reactive antibody response with no detectable production of antibodies that target novel epitopes on the RBD of the BA.5 spike.

**Figure 3.**
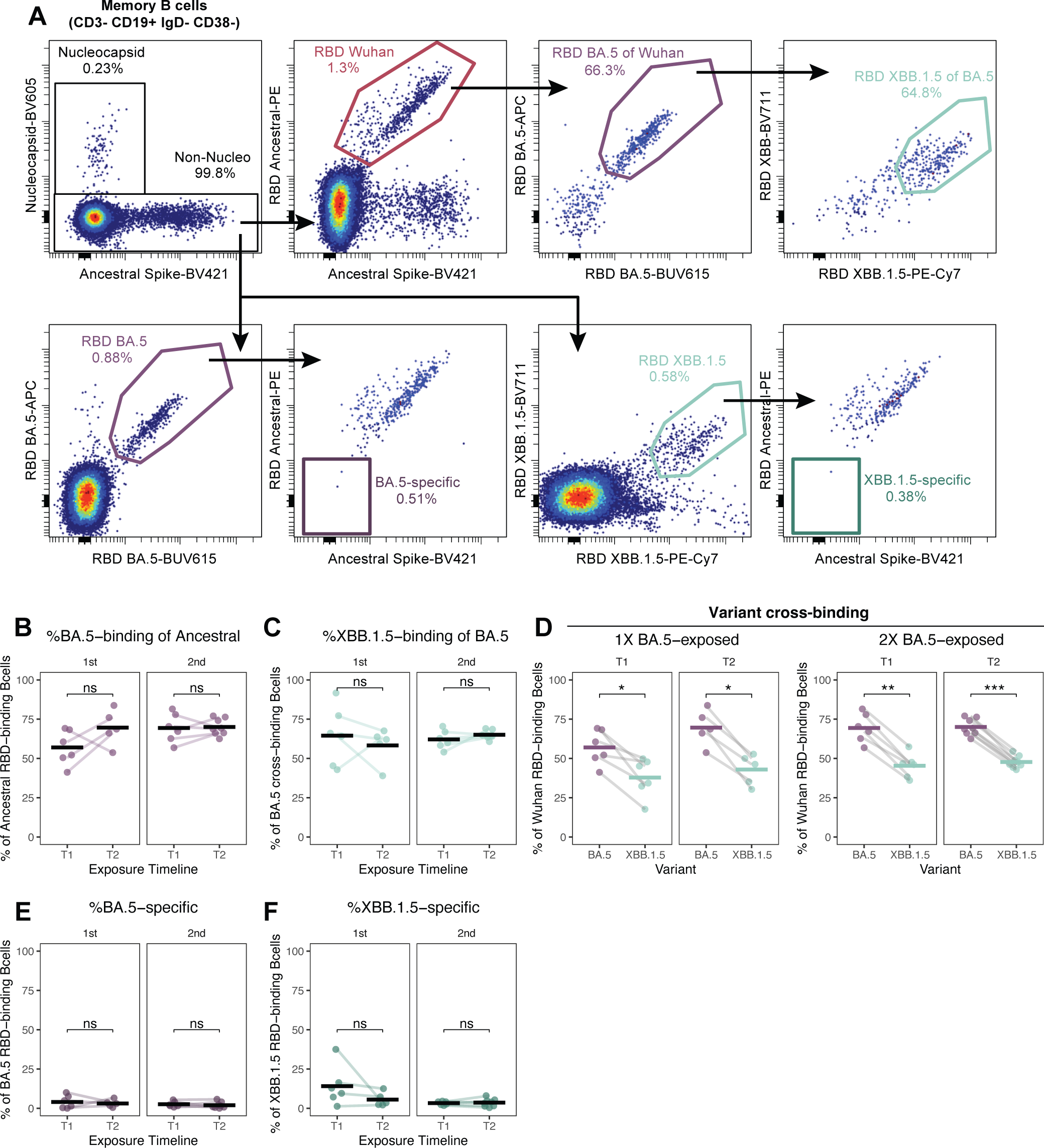
Cross-reactive B cells are recruited in response to primary and secondary BA.5 exposures in vaccinated individuals. A) Representative flow-cytometry plot showing gating strategy for probe positive memory B cells. B) The percent of ancestral RBD-binding B cells that cross-bind to BA.5 RBD before and after first (left) and second (right) exposure to BA.5. C) The percent of ancestral RBD and BA.5 RBD cross-binding B cells that also cross-bind to XBB.1.5 RBD before and after first (left) and second (right) exposure to BA.5. D) The percent of ancestral RBD-binding B cells that cross-bind BA.5 and XBB.1.5 RBD before and after first (left) and second (right) exposure to BA.5. E-F) The frequency of BA.5 RBD-binding (E) and XBB.1.5 RBD-binding (F) B cells that do not cross-react with ancestral RBD before and after first (left) and second (right) exposure to BA.5.

### BA.5 exposures elicit antibodies targeting conserved RBD residues mutated in XBB.1.5

Our data suggest that BA.5 exposures boost antibodies that target spike residues that are conserved between ancestral and BA.5 variants but are mutated in XBB.1.5. To map the specificity of these antibodies, we completed additional absorption assays with BA.5 spike proteins that were engineered to possess different RBD amino acid substitutions. We absorbed serum samples with BA.5 spike mutants that had one or two XBB.1.5 RBD amino acid substitutions at residues that are shared between ancestral and BA.5 spikes but differ in XBB.1.5 (spike amino acid residues 346, 368, 445/446, 460, and 490) (**Figure 4A**). We also absorbed serum with 7 additional BA.5 spike proteins with different combinations of XBB.1.5 RBD mutations, including a mutant that possessed all RBD substitutions that are shared between ancestral and BA.5 spikes but differ in XBB.1.5 spike (protein M7, **Figure 4B**). Absorption with BA.5 spikes with each single substitution decreased the ability of serum antibodies to bind wildtype BA.5, suggesting that a fraction of polyclonal antibodies targeted each mutated site (**Figure 4C**). Absorption with BA.5 proteins with multiple mutations further decreased the binding of BA.5-reactive antibodies (**Figure 4C**). Similar results were obtained when we repeated experiments with sera isolated from ancestral vaccinated individuals after two BA.5 exposures (**Figure 4D**). Sera absorbed with the M7 protein containing all XBB.1.5 RBD substitutions efficiently neutralized BA.5, indicating that a large fraction of BA.5 neutralizing antibodies targeted epitopes that are conserved between the ancestral and BA.5 RBDs but not the XBB.1.5 RBD (**Figure 4E, F, S4**). Collectively, these data support the hypothesis that BA.5 exposures elicit antibodies that target epitopes conserved in the ancestral spike RBD that later became mutated in the XBB.1.5 variant.

**Figure 4.**
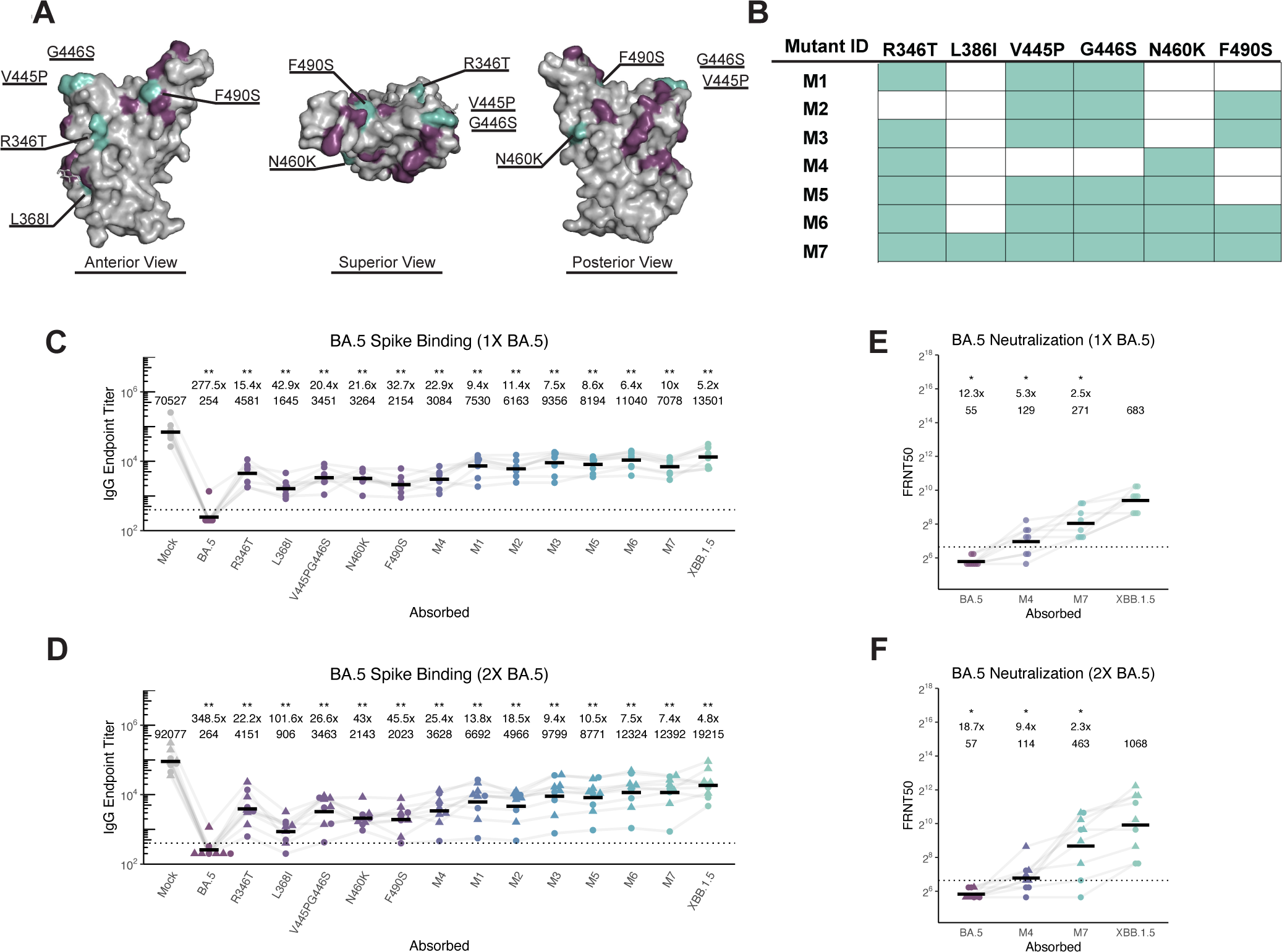
Antibodies elicited by successive ancestral SARS-CoV-2 and BA.5 exposures target conserved residues that are mutated in the XBB.1.5 RBD. A) Ancestral RBD structure with residues that are different in BA.5 are highlighted in purple, and residues conserved between ancestral and BA.5 but different in XBB.1.5 are highlighted in teal (PDB 6M0J). B) List of recombinant mutations created for this study using the BA.5 spike backbone. C) IgG ELISA using T2 breakthrough sera after absorption with single mutants and those listed in panel B. D) IgG ELISA usingT2 sera from individuals exposed twice with BA.5 after absorption with single mutants and those listed in panel B. E,F) Neutralization assays were performed using T2 absorbed serum from single (E) and two (F) BA.5 exposures using BA.5 pseudotype virus. For all, individual points are average of n = 2 technical replicates. Black bars indicate geometric mean. Wilcoxon signed-rank test with benjamini-hochberg correction for multiple testing. All comparisons to mock absorption. * p<0.05, ** p<0.01.

### XBB exposures elicit antibodies cross-reactive to the ancestral spike in individuals previously vaccinated with mRNA-LNP vaccines expressing the ancestral spike

XBB, the recombinant variant of BA.2.10.1 and BA.2.75 sublineages containing ∼43 spike mutations, and its subvariants, namely XBB.1.5, have dominated infections since late 2022 and resulted in a shift to a new monovalent booster vaccine in September 2023 (CDC, Selvavinayagam et al., Wang et al., 2023; WHO 2022). We longitudinally sampled individuals who were exposed to XBB.1.5 via the monovalent mRNA-LNP booster (n = 12) or by XBB-subvariant breakthrough infections (symptom onset between 1/2/2022 and 10/1/2022, n = 10) and assessed the specificity and functionality of antibody and B cell responses (**Figure 5A**). We observed a robust increase in antibodies that bound and neutralized the ancestral and XBB.1.5 variant 15 and 45 days after XBB.1.5 monovalent mRNA-LNP vaccination or XBB.1.5 infection (**Figure 5B-D**). While increases in antibody titers to both viruses were significant, the fold-increase in XBB.1.5 neutralizing titers was ∼5-fold higher than that of ancestral strain, and neutralization potency of XBB.1.5 antibodies increased slightly following XBB.1.5 exposure, whereas that of ancestral did not (**Figure 5D-E**). We found that XBB.1.5 monovalent mRNA-LNP vaccination rapidly boosted XBB.1.5 neutralizing antibodies that peaked 15 days after vaccination, whereas antibodies elicited by XBB.1.5 infections steadily rose for 45 days after exposure, consistent with earlier reports of slower kinetics of immune responses elicited by breakthrough infections (Koutsakos et al., 2022; Painter et al., 2023) (**Figure S5A-C)**. Although early kinetics differed, antibodies elicited by XBB.1.5 infections and vaccinations were at similar levels by 45 days after exposure (**Figure S5A-C**).

**Figure 5.**
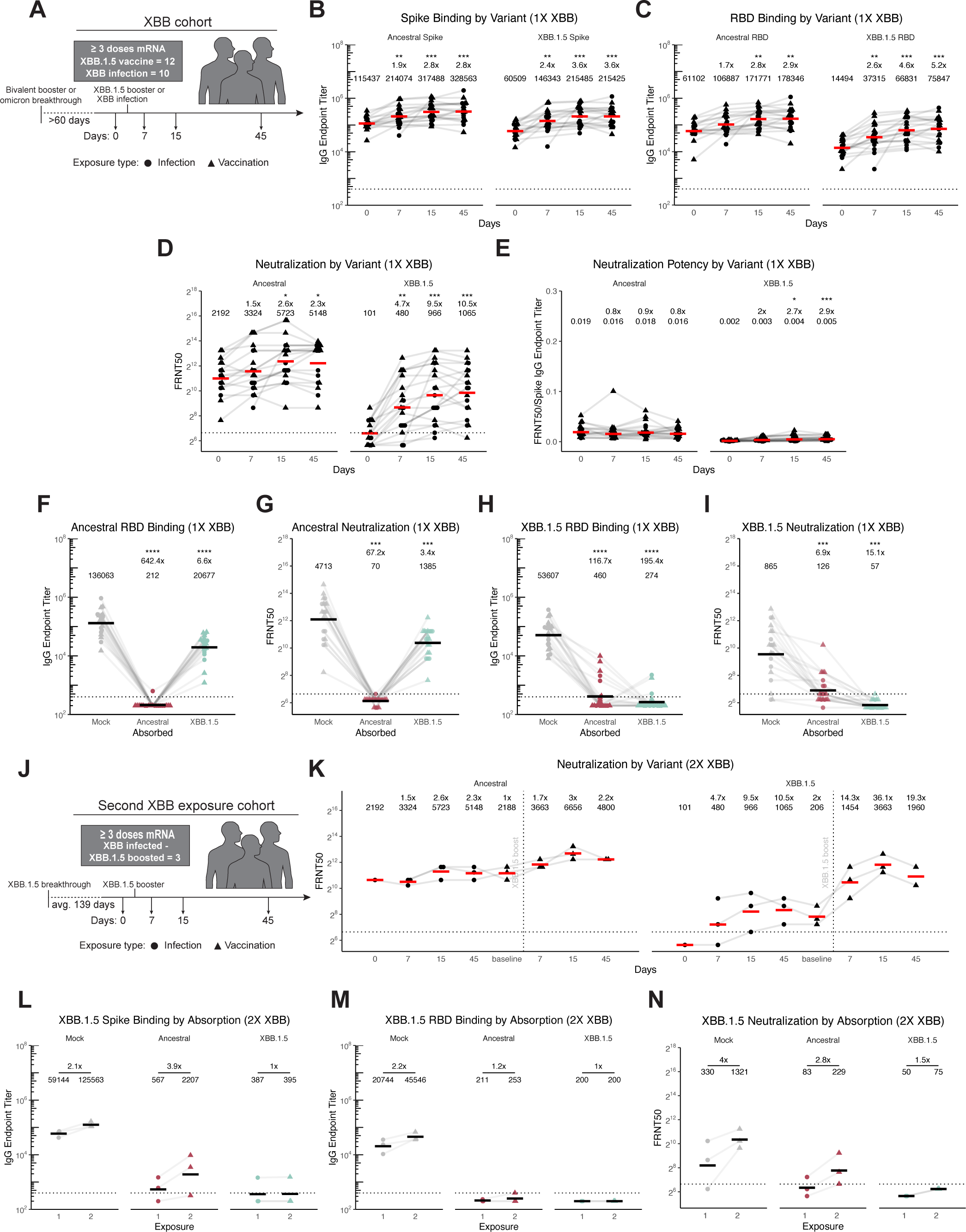
Primary and secondary XBB exposures elicit mostly antibodies that cross-react to the ancestral SARS-CoV-2 spike. A) Schematic of participants in this study who were exposed to XBB. B)-C) Antigen-specific IgG ELISAs were performed using sera from timepoints indicated in panel A) against ancestral and XBB.1.5 full-length spike (B) and RBD (C). Endpoint titers are reported as reciprocal dilutions. D) Neutralization assays were performed using serum obtained at timepoints indicated in panel A) against ancestral and XBB.1.5 SARS-CoV-2 pseudoviruses. Values reported are focus reduction neutralization test (FRNT) 50, or reciprocal serum dilution at which <50% viral input foci are observed. E) Neutralization potency was calculated by dividing FRNT50 values by spike IgG titer. F),H) Antigen-specific IgG ELISAs were performed using sera (collected 45 days after XBB1.5 exposure) after absorption with ancestral or XBB.1.5 spike proteins. G),I) SARS-CoV-2 pseudotype neutralization assays were performed using sera (collected 45 days after XBB.1.5 exposure) after absorption with ancestral or XBB.1.5 spike proteins. J) Schematic of participants in this study who were exposed twice to XBB. K) SARS-CoV-2 pseudotype neutralization assays were performed using sera obtained at timepoints indicated in panel J) using ancestral and XBB.1.5 pseudoviruses. L),M) Antigen-specific IgG ELISAs and N) SARS-CoV-2 neutralization assays were performed using sera (collected 45 days after each XBB.1.5 exposure) after absorption with ancestral or XBB1.5 spike proteins. For all, Individual points are average of n = 2 technical replicates. Red/black bars indicate geometric mean. Wilcoxon rank-sum test with benjamini-hochberg correction for multiple testing. All comparisons to timepoint 1 or mock absorption. * p<0.05, ** p<0.01, *** p<0.001, **** p<0.0001.

We performed absorption assays using sera collected 45 days after XBB.1.5 exposures to identify antibody specificities elicited by XBB infections and monovalent vaccinations. Beads coated with the XBB.1.5 spike poorly absorbed antibodies that bound (**Figure 5F**) and neutralized (**Figure 5G**) the ancestral SARS-CoV-2 strain. Conversely, beads coated with the ancestral spike absorbed most XBB.1.5-reactive and neutralizing antibodies (**Figure 5H-I, S5E**), suggesting that the majority of antibodies elicited by XBB exposures bound to epitopes conserved in the ancestral spike. While most individuals produced XBB.1.5-reactive antibodies that targeted epitopes conserved in the ancestral spike, we identified some individuals who produced XBB.1.5-specific responses that were only partially absorbed by beads coated with the ancestral spike (**Figure 5H-I, S5E**). These data suggest that XBB.1.5, which is more antigenically distant compared to BA.5, is capable of eliciting variant-specific responses in a subset of individuals who were previously vaccinated with mRNA-LNP expressing the ancestral spike.

Next, we characterized antibodies in sera from 3 individuals who were infected with an XBB-variant and subsequently received the XBB.1.5 monovalent booster (average 139 days after infection, **Figure 5J**). Unlike a second BA.5 exposure, a second XBB exposure greatly increased XBB.1.5 neutralizing antibody titers and potency (**Figure 5K, S5F-H)**. Using absorption assays, we also observed increases in both XBB.1.5-specific binding and neutralizing antibodies following the second XBB exposure (**Figure 5L-N, S5I-K)**. Similar to single XBB exposures, the majority of neutralizing antibodies elicited by two XBB exposures targeted epitopes in the ancestral spike, with XBB.1.5-specific antibodies constituting only 17% of total XBB.1.5 neutralizing antibodies. Nonetheless, these experiments demonstrate that XBB can provoke variant-specific responses in a subset of individuals who previously received mRNA-LNP vaccines expressing the ancestral spike, and that these responses can be further boosted by sequential XBB exposures.

### Cross-reactive B cell responses dominate following XBB exposures in individuals previously vaccinated with mRNA-LNP vaccines expressing the ancestral spike

To further explore the relationships between cross-reactive and variant-specific responses elicited by XBB, we analyzed antigen-specific B cells elicited after XBB exposures. We observed a significant increase in ancestral and XBB.1.5-reactive B cells by 15 days post-exposure, followed by a slight contraction by day 45 (**Fig 6A-B)**. The percent of ancestral RBD-binding B cells that were cross-reactive with XBB.1.5 increased significantly by day 7 and remained above baseline at day 45 (**Fig. 6C**). The frequencies of XBB.1.5-specific B cells (cells that did not cross-react with ancestral RBD) were low but significantly expanded between days 0 and 7 before contracting by day 45 (**Fig. 6D, Fig. S6A**). There was substantial heterogeneity in the magnitude of XBB.1.5-specific B cell expansion, suggesting that variant-specific responses may be restrained in some individuals (**Fig. 6D**). Despite expanding significantly during acute XBB exposures, the proportion of XBB.1.5 RBD-binding B cells that were specific for XBB.1.5 did not increase, consistent with the cross-reactive response remaining dominant compared to variant-specific responses (**Fig. 6E**).

**Figure 6.**
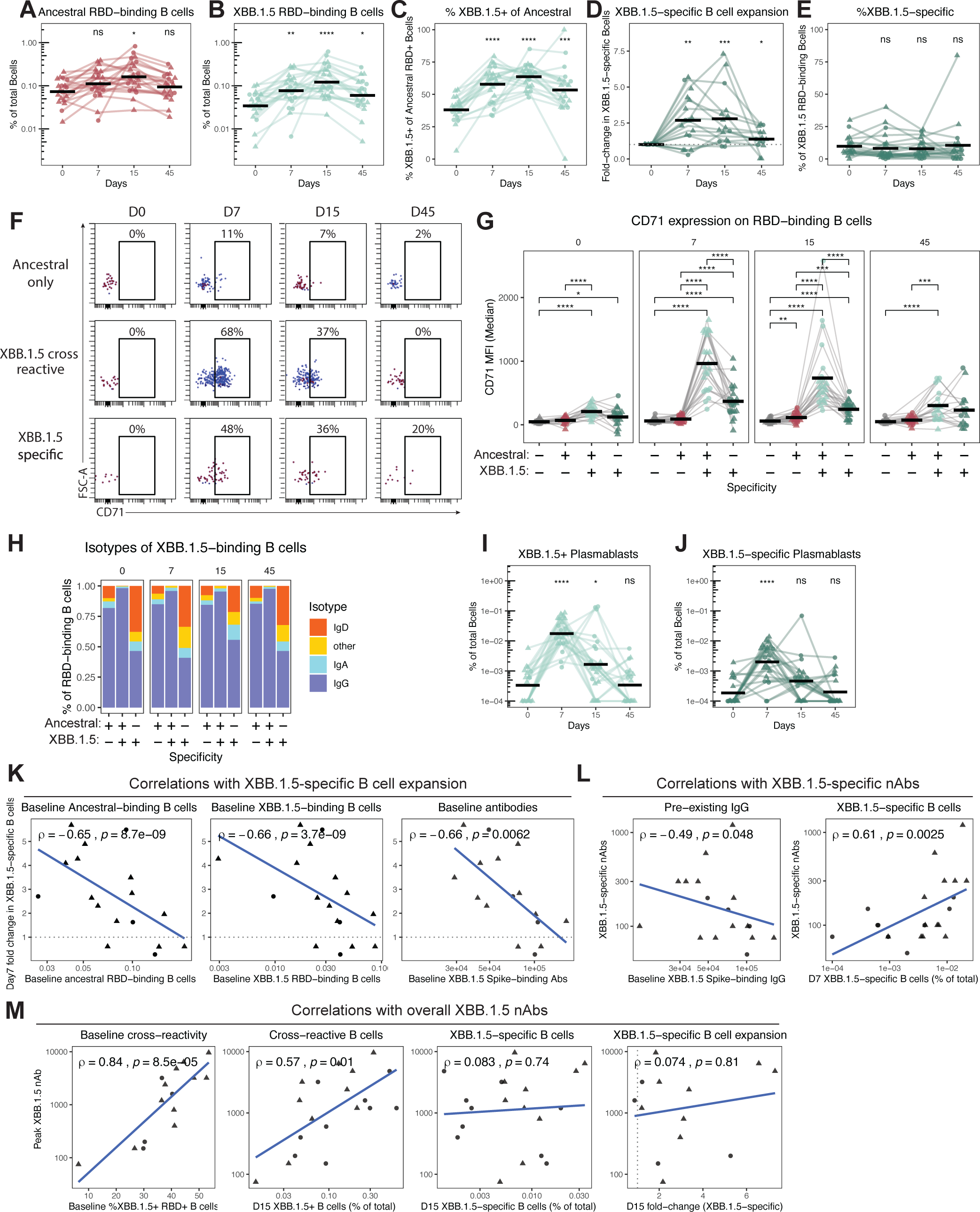
Cross-reactive B cells dominate immune responses elicited by XBB, but variant-specific responses are observed in individuals with low pre-existing immunity. A-B) The percent of total B cells that bind A) ancestral RBD and B) XBB.1.5 RBD before and after exposure to XBB.1.5. C) The percent of ancestral RBD-binding B cells that cross-bind to XBB.1.5 RBD before and after exposure to XBB.1.5. D) The fold expansion of B cells that bind XBB.1.5 RBD but not ancestral RBD (XBB.1.5-specific) following XBB.1.5 exposure. E) The percent of total XBB.1.5 RBD-binding B cells that do not bind ancestral RBD. F) Representative flow cytometry plots depicting longitudinal changes in CD71 expression on different RBD-binding B cell populations from a single individual. G) Summary data of median CD71 expression on the indicated RBD-binding B cell populations at 0, 7, 15, and 45 days post-XBB exposure. H) Summary data of the isotype distribution of RBD-binding B cells at 0, 7, 15, and 45 days post-XBB exposure. I-J) The percent of total B cells that are CD38+CD27+ plasmablasts and I) bind XBB.1.5 RBD or J) bind XBB.1.5 RBD and do not bind ancestral RBD (XBB.1.5-specific). K-M) Correlations within B cell and antibody responses. K) Correlation of day 7 fold change in XBB.1.5-specific B cells as in panel D with day 0 percent of total B cells that bind ancestral RBD (left); day 0 percent of total B cells that bind XBB.1.5 RBD (center); day 0 XBB.1.5 spike-binding antibodies by ELISA (right). L) Correlations of ancestral spike absorbed XBB.1.5 neutralizing antibody titers at day 45 (as in Fig. 5N) with day 0 XBB.1.5 spike-binding antibodies quantified by ELISA (left), and the percent of total B cells that bind XBB.1.5 RBD but not ancestral RBD at day 7 (right). M) Correlation of overall day 45 XBB.1.5 neutralizing antibody titers (unabsorbed) with the day 0 percent of ancestral RBD+ B cells that cross-bind XBB.1.5 RBD; the day 15 percentage of total B cells that bind XBB.1.5 RBD, the day 15 percentage of total B cells that bind XBB.1.5 RBD but do not bind ancestral RBD (XB.1.5-specific), and the day 15 fold change in XBB-specific B cells as in D. Points represent individual subjects and thin lines indicate individual subjects sampled longitudinally. Horizontal bars represent means. Statistics were calculated using two-sided Wilcoxon test with Benjamini-Hochberg correction for multiple comparisons. Statistics without brackets are in comparison to day 0. Correlation statistics were calculated using Spearman rank correlation and are shown with Pearson trend lines for visualization.

CD71 is expressed following B cell activation, with reactivated memory B cells expressing higher CD71 than stimulated naïve B cells (Auladell et al., 2019). CD71 was rapidly upregulated after XBB exposure and was significantly elevated on XBB.1.5 cross-reactive and XBB.1.5-specific B cells at day 7 and day 15 compared to bulk B cells or B cells that bound the ancestral RBD but not the XBB.1.5 RBD (**Fig. 6F-G, S6B**). XBB.1.5-specific B cells expressed lower CD71 than XBB.1.5 cross-reactive B cells, consistent with differences between recall responses from memory B cells and *de novo* responses arising from naïve B cells (Auladell et al., 2019). XBB.1.5 cross-reactive B cells were class-switched and expressed IgG, whereas most XBB.1.5-specific B cells were not class-switched to IgA or IgG (**Fig. 6H**), suggesting that they were likely derived from *de novo* responses. In addition to rapid CD71 upregulation, XBB.1.5-binding plasmablasts, including XBB.1.5-specific plasmablasts, increased dramatically by day 7 after XBB.1.5 exposure (**Fig 6I-J**). Taken together, these data suggest that XBB elicits a potent B cell response that is largely cross-reactive with the ancestral spike, with some individuals generating variant-specific B cells.

Finally, we wanted to determine what cellular or serological metrics were associated with the production of XBB.1.5-specific responses. Elevated baseline antibody titers, ancestral and XBB.1.5 RBD-binding B cell frequencies, and XBB.1.5 cross-reactive B cell frequencies were all associated with diminished XBB.1.5-specific B cell responses (**Fig. 6K, S6C-D**). Moreover, high pre-existing antibody titers against XBB.1.5 spike were associated with decreased XBB.1.5-specific neutralizing antibody titers (**Fig. 6L**). As expected, there was a positive correlation between XBB.1.5-specific B cells at day 7 and peak XBB.1.5-specific neutralizing antibody titers (**Fig. 6L**). XBB.1.5-specific antibodies were also correlated with XBB.1.5-specific B cell expansion and the percent of XBB.1.5-binding B cells that were XBB.1.5-specific (**Fig. S6E**). Together, these analyses support a model in which pre-existing B cells and antibodies influence the development of variant-specific responses, where individuals with lower pre-existing antibody titers and fewer cross-reactive memory B cells are better able to activate naïve XBB.1.5-specific B cells and produce XBB.1.5-specific antibodies. However, baseline cross-reactive B cells are strongly correlated with peak XBB.1.5 neutralizing antibody titers, whereas XBB.1.5-specific B cell responses have no association with overall XBB.1.5 neutralizing antibody titers after infection or vaccination (**Fig. 6M**). This is consistent with the observation that the majority of XBB.1.5 neutralizing antibodies cross-react to the ancestral spike. Overall, these analyses highlight the diversity of the humoral response to XBB and define several key factors contributing to the development of variant-specific B cells and antibodies.

## Discussion

Through longitudinal sampling and intensive serological and cellular analyses, our studies demonstrate that B cells elicited by ancestral SARS-CoV-2 mRNA vaccines are efficiently recalled by SARS-CoV-2 variants, leading to the rapid production of neutralizing antibodies against epitopes conserved between ancestral and variant strains. We found that the BA.5 spike elicits antibodies that recognize epitopes conserved between the ancestral and BA.5 spikes, but fail to bind to the more antigenically distant XBB.1.5 spike. Similarly, the majority of antibodies elicited by XBB infection or monovalent vaccination targeted epitopes conserved between the ancestral and XBB.1.5 spikes. Both BA.5 and XBB elicited responses that were dominated by memory B cells primed by prior ancestral mRNA-LNP vaccinations; however, unlike BA.5, we found that XBB elicited low levels of variant-specific B cell and antibody responses in some individuals.

Our data are consistent with reports showing antibody responses to SARS-CoV-2 variants typically cross-react to the ancestral spike in individuals who initially encountered ancestral SARS-CoV-2 antigens (Addetia et al., Chalkias et al., 2023; Kared et al., Koutsakos et al., 2022; Painter et al., 2023; Park et al., 2022; Tortorici et al., 2023; Wang et al., 2022; Wang et al., Weber et al., Yisimayi et al., 2023). Clinical studies have shown that BA.5 and XBB.1.5 variant booster vaccines are highly effective at reducing deaths and hospitalizations caused by circulating variants (Carr et al., Hoffmann et al., Lin et al., Lin et al., Shresta et al., Tan et al., Tan et al., Wang et al., 2023). It is likely that antibodies targeting epitopes conserved with the ancestral SARS-CoV-2 spike contribute to variant booster vaccine protection. It is important to highlight that cross-reactive B cells elicited by earlier ancestral SARS-CoV-2 mRNA-LNP vaccinations are strongly correlated with the induction of high levels of XBB.1.5 neutralizing antibodies after monovalent XBB.1.5 vaccination. Therefore, while ‘immune imprinting’ by ancestral SARS-CoV-2 mRNA-LNP vaccines clearly influence the specificity of antibodies elicited by variant infections and vaccinations, these prior vaccinations are also highly beneficial for establishing memory B cells that can be rapidly recruited to produce neutralizing antibodies against SARS-CoV-2 variants.

Further studies should evaluate if SARS-CoV-2 antigens with greater antigenic distances are able to efficiently elicit variant-specific *de novo* antibody responses in individuals who were previously vaccinated with ancestral mRNA-LNP vaccines. In our studies, XBB exposures promoted the development of low-level variant-specific XBB.1.5-specific antibody responses in some individuals. These variant-specific responses were associated with lower baseline SARS-CoV-2 XBB.1.5-specific antibody titers and B cell frequencies, suggesting that factors such as epitope masking and feedback inhibition may potentially limit the recruitment of naïve B cells that target novel epitopes on variant spike proteins (Bergström et al., 2017; Schaefer-Babajew et al., 2023; Tas et al., 2022). It will also be important to quantify the level of somatic hypermutation in B cells that produce antibodies that bind specifically to variants. While it is likely that variant-specific antibodies in our studies are derived from *de novo* activated B cells, it is also possible that these antibodies are produced from memory B cells that have lost binding to ancestral spike through somatic hypermutation.

Future studies should also determine if mRNA-LNP and inactivated virus vaccines elicit different ‘immunological imprints’ that affect the specificity and magnitude of antibody responses elicited by variant infections and vaccinations. Comparative analyses have previously focused on general immunogenicity and effectiveness against severe disease, showing higher antibody titers and better overall protection in mRNA-LNP vaccinated individuals compared to inactivated virus vaccinated individuals (Lim et al., 2021, Premikha et al., 2019), Yisimayi and colleagues recently demonstrated that individuals who previously received inactivated SARS-CoV-2 vaccine produced robust variant-specific responses after subsequent Omicron exposures (Yisimayi et al., 2023). Taken with our data and others, this suggests that inactivated virus vaccines potentially establish weaker ‘immunological imprints’ compared to mRNA-LNP vaccines.

The human immune landscape against SARS-CoV-2 is becoming more heterogenous as variants emerge and infection and vaccination histories become diverse among different individuals. Most humans have been ‘immunologically imprinted’ with antigens from the ancestral SARS-CoV-2 strain, but that will inevitably change as time progresses. Most children born today will be first introduced to SARS-CoV-2 antigens in the form of a variant infection or variant vaccination, leading to the formation of different memory B cell populations compared to individuals first exposed to ancestral SARS-CoV-2 antigens. Though the impact of birth year imprinting on the evolution of future SARS-CoV-2 variants is unknown, there is evidence of influenza epidemic virus susceptibilities being shaped by childhood exposures (Arevalo et al., 2020; Gostic et al., 2016; Gostic et al., 2019). Overall, an improved understanding of how SARS-CoV-2 immune history influences antibody specificity to new variants will be critical for rationally designing future vaccines and mitigating disease burden.

### Limitations of this study

While most individuals in these cohorts followed a similar 3-dose vaccination with mRNA-LNP expressing the ancestral spike followed by BA.5 breakthrough infection, some individuals experienced infections with prior variants, as shown in **Supplemental Table 1**. The XBB cohort included individuals with a more diverse exposure history. We did not follow a single longitudinal cohort of individuals following first and second BA.5 exposures and through subsequent XBB.1.5 vaccinations, which introduces variance between all cohorts. Our antibody functionality assays were limited to pseudovirus neutralization assays, which have been shown to be highly correlative with live virus neutralization assays. For our BA.5 cohorts, we only measured functionality of antibodies at two timepoints and specificity at a single timepoint; it is possible epitope targeting continues to change over time after antigen exposure. Additional analyses of late timepoints will be important to include in future studies to understand how antibody specificities change over time.

## Data Availability

All data produced in the present work are contained in the manuscript

## Acknowledgements

We would like to thank the study participants for their generosity in making the study possible. We also thank the Penn Cytomics and Cell Sorting Resource Laboratory for access to instruments, members of the Hensley and Wherry labs for helpful discussions and feedback, and the Immune Health team. This project has been funded in part with Federal funds from the National Institute of Allergy and Infectious Diseases, National Institutes of Health, Department of Health and Human Services, under Contract No. 75N93021C00015 (S.E.H. and E.J.W.) and Grant Nos. U19AI082630 (S.E.H. and E.J.W.), AI105343 (E.J.W.), AI108545 (E.J.W.), AI155577 (E.J.W.), AI149680 (E.J.W.); and the Parker Institute for Cancer Immunotherapy (to EJW). S.E.H. holds an Investigators in the Pathogenesis of Infectious Disease Awards from the Burroughs Wellcome Fund. D.B.R. was supported by an MD fellowship of the Boehringer Ingelheim Fonds.

## Author Contributions

Conceptualization, T.S.J., M.M.P., E.J.W., and S.E.H.; Methodology, T.S.J., S.H.L., and M.M.P.; Investigation, T.S.J., S.H.L., R.K.A., N.R.D., D.B.R., and M.M.P.; Formal Analysis, T.S.J. and M.M.P.; Visualization, T.S.J. and M.M.P.; Writing – Original Draft, T.S.J. and S.E.H., Writing – Review & Editing, T.S.J., S.H.L., M.M.P., E.J.W., D.C.D., and S.E.H.; Supervision, D.C.D, E.J.W., and S.E.H.; Funding Acquisition, E.J.W. and S.E.H.; Resources, E.J.W. and S.E.H.

## Declaration of Interests

E.J.W. is a member of the Parker Institute for Cancer Immunotherapy. E.J.W. is an advisor for Arsenal Biosciences, Coherus, Danger Bio, IpiNovyx, Janssen, New Limit, Marengo, Pluto Immunotherapeutics Related Sciences, Santa Ana Bio, and Synthekine. E.J.W. is a founder of and holds stock in Coherus, Danger Bio, and Arsenal Biosciences.

## Inclusion and Diversity

We support inclusive, diverse, and equitable conduct of research.

## Methods

### Human Subject Recruitment and Sampling

Human subjects were recruited for this study for longitudinal sampling before and after either mRNA booster vaccination or SARS-CoV-2 breakthrough infection (IRB#851465). Full cohort and demographic information are provided in **Supplemental Table 1**. Additional healthy donor PBMC samples were collected with approval from the University of Pennsylvania Institutional Review Board (IRB# 845061). For many individuals, baseline samples were acquired prior to SARS-CoV-2 infection or vaccination but at least 2 months after their most recent infection or vaccine dose. These samples are labeled as day 0 throughout the manuscript. Samples were collected on days 7, 15, and 45 based on the date of booster vaccination or the reported date of first symptom onset (IRB#851465). Prior SARS-CoV-2 infection status was determined by self-reporting. All participants were otherwise healthy, with no self-reported history of chronic health conditions, and none were hospitalized during SARS-CoV-2 infection. Peripheral blood samples (30-100mL) and clinical questionnaire data were collected at each study visit.

### Peripheral blood sample processing

Following standard phlebotomy procedures, 30-100mL of venous blood was collected into sodium heparin tubes. Plasma was separated by centrifugation at 1800xg for 15 minutes, aliquoted, stored at −80°C, and heat inactivated for 30 minutes at 56°C prior to binding and neutralizing antibody analyses. After removing plasma, the remaining fractions were diluted with RPMI + 1% FBS + 2mM L-Glutamine + 100 U Penicillin/Streptomycin (R1) to achieve a final volume double that of the original whole blood. The diluted blood was then layered over 15mL lymphoprep gradients (STEMCELL Technologies) in SEPMATE tubes (STEMCELL Technologies) and spun at 1200g for 10 minutes. The peripheral blood mononuclear cell (PBMC) fraction was harvested from SEPMATE tubes, washed once with R1 and pelleted, and cell pellets were resuspended in ACK lysis buffer (Thermo Fisher). After 5 minutes of red blood cell lysis at room temperature, the lysis was quenched by adding R1. PBMCs were then filtered through a 70μm cell strainer and counted using a Countess automated cell counter (Thermo Fisher). Aliquots containing 10×10^6^ PBMCs were cryopreserved in 90% FBS 10% DMSO and stored at −80°C for later flow cytometric analyses.

### Recombinant Protein Production and Expression

SARS-CoV-2 full-length spike or RBD plasmids were transfected into 293F cells at 1e6 cells/mL at a 1µg :1mL ratio. Cells were incubated for 6 days (full-length spike) or 4 days (RBD) at 37°C and 150 RPM before purification. To purify, cultures were centrifuged at 3200xg for 6 minutes to clarify supernatant. Ni-NTA resin (Qiagen) was resuspended and 5mL was transferred into a 50mL conical for each flask being purified. Volume was brought to 50mL with sterile PBS and conical tubes were centrifuged for 10 minutes at 3200xg. After centrifugation, excess PBS was removed, and resin was added to clarified supernatant. Resin-supernatant mixtures were incubated for 2 hours at 4°C and 220 RPM. After incubation, resin-supernatant was added to gravity columns (Bio-rad), then washed 4X with wash buffer (50 mM NaHCO3, 300 mM NaCl, 20 mM imidazole, pH 8) and eluted in a total volume of 8 mL elution buffer (50 mM NaHCO3, 300 mM NaCl, 300 mM imidazole, pH 8). Eluted protein was concentrated and PBS buffer exchanged using 10kDa (RBD) or 30kDa (full-length spike) centrifugal filters (Sigma-Aldrich). Protein concentrations were measured by BCA assay (ThermoFisher).

### Carboxyl-Magnetic Bead Coupling and Serum Absorptions

Recombinant spike proteins were coupled to carboxyl magnetic beads (Ray-biotech) at a ratio of 35 µg antigen to 100 µL beads. The bead-antigen mixture was vortexed then incubated at 4°C with rocking for 2 hours. The unbound fraction was removed by placing beads to a magnetic stand, and beads were quenched by incubating in 50 mM Tris, pH 7.4 for 15 minutes at RT with rocking. After 15 minutes, beads were placed back on the magnet to remove the quenching buffer. Beads were washed 4x with wash buffer (DPBS supplemented with 0.1% BSA and 0.05% Tween-20). For each wash, 1 bead volume of wash buffer was added to the beads, vortexed, then placed back on the magnet. Beads were finally resuspended in 1 bead-volume of wash buffer before storage at 4°C.

For absorptions, sera were first diluted 1:35.7 with sterile DPBS and then brought to 1:50 dilution with the addition of recombinant protein-coupled beads at a final bead:sera volume ratio of 1:2. The bead-sera mixture was vortexed, then shaken at 1100 RPM for 1 hour at RT. Bead-sera mixtures were then placed on a magnet for separation and unabsorbed fractions were removed and transferred to a clean tube.

### Antigen-specific ELISA

Antigen-specific ELISAs were performed as previously described (Anderson et al., 2021). Briefly, high-binding plates (ThermoFisher) were coated with 2 µg/mL of recombinant protein or with DPBS to control for background antibody binding at 4°C overnight. The next day, plates were washed 3x with PBS containing 0.1% Tween 20 (PBS-T) and blocked for 1 hour with 200µL blocking buffer (DPBS supplemented with 3% milk powder and 0.1% tween-20). Plates were then washed 3x with PBS-T, and 50µL serum diluted in dilution buffer (DPBS supplemented with 1% milk and 0.1%Tween-20) were added. After 2 hours of incubation, plates were washed 3x with PBS-T and 50 µL goat anti-human IgG HRP conjugate (Jackson) was added to each well and allowed to incubate for 1 hour. Plates were washed 3x with PBS-T, then 50 µL SureBlue 3,3’,5,5’-tetramethylbenzidine substrate (SeraCare) was added to each well. The reaction was allowed to incubate in the dark for 5 minutes before addition of 25 µL 250 mM hydrochloric acid stop solution. Plates were shaken to distribute solution and read at OD450nm immediately using a Spectramax plate reader (Molecular Devices). Serum antibody titers were obtained from a standard curve of serially diluted pooled serum (starting dilution 1:100). Standard curves were included on all plates for plate-to-plate variation. PBS controls were included for each sample dilution plate to account for nonspecific binding. Antibody titers for each sample were measured in two technical replicates performed on separate days.

### SARS-CoV-2 Pseudovirus Production

SARS-CoV-2 pseudotyped vesicular stomatitis virus (VSV) were produced as previously described (Anderson et al., 2021; Goel et al., 2021). Briefly, 293T cells were seeded at 3.5×10^6^ cells in collagen-coated 10 cm tissue culture dish, incubated for 24 hours, and transfected using calcium phosphate with 25 μg of SARS-CoV-2 variant plasmids encoding a codon optimized SARS-CoV-2 spike gene with an 18-residue truncation in the cytoplasmic tail. After 24 hours, the SARS-CoV-2 Spike-expressing cells were infected for 2-4 hours with VSV-G pseudotyped VSVΔG-RFP at an MOI of ∼2-4. Virus-containing media was removed, and the cells were re-fed with media. VSVΔG-RFP SARS-CoV-2 pseudotypes were harvested 24-28 hours after infection, clarified by centrifugation twice at 6000xg, then aliquoted and stored at −80°C.

### Quantification of SARS-CoV-2 pseudotype neutralizing antibody titers

Focus reduction neutralization titers were determined as previously described. Briefly, 96-well collagen-coated plates were seeded with 2.5 x 10^4^ TMPRSS2 expressing VeroE6 cells per well the night prior. The next day, heat-inactivated sera or post-absorption sera were serially diluted 2-fold and mixed with 200-300 focus forming units per well of VSVΔG-RFP SARS-CoV-2 pseudotype viruses. Media used for serum and virus dilution contained 600 ng/mL of an anti-VSV-G antibody 1E9F9 to ensure neutralization of any VSV-G carryover virus from pseudotype production. The serum-virus mixture was incubated for 1 hour at 37°C before being plated on VeroE6 TMPRSS2 cells. After 22 hours of incubation at 37°C, plates were washed with DPBS and fixed with 4% paraformaldehyde. The spots were visualized and counted on an S6 FluoroSpot Analyzer (CTL). The focus reduction neutralization titer 50% (FRNT50) was determined as the greatest serum dilution at which focus count was reduced by at least 50% relative to control wells infected with only pseudotype viruses in the absence of human serum. FRNT50 titers for each sample were measured in two technical.

### SARS-CoV-2-specific B cell flow cytometric analyses

Antigen-specific B cells were detected using biotinylated proteins in combination with different streptavidin (SA)–fluorophore conjugates as described (Goel et al., 2021). All reagents are listed in **Supplemental Table 2**. For each sample, biotinylated proteins were multimerized with fluorescently labeled streptavidin (SA) for 1.5 h at 4 °C at the following ratios, all of which are ∼4:1 molar ratios calculated relative to the SA-only component irrespective of fluorophore: 200 ng full-length spike protein was mixed with 20 ng SA-BV421, 30 ng NTD was mixed with 12 ng SA-BV786, 25 ng WT RBD was mixed with 12.5 ng SA-PE, 25 ng XBB.1.5 RBD was mixed with 12.5 ng SA-BV711, 25 ng XBB.1.5 RBD was mixed with 12.5 ng SA-PE-Cy7, 25 ng BA.5 RBD was mixed with 12.5 ng SA-BUV615, 25 ng BA.5 was mixed with 12.5 ng SA-APC, 50 ng S2 was mixed with 12 ng SA-BUV737, and 50 ng nucleocapsid protein was mixed with 14 ng SA-BV605. In total, 12.5 ng SA-PE-Cy5 was used as a decoy probe without biotinylated protein to gate out cells that nonspecifically bound SA. All experimental steps were performed in a 50/50 mixture of PBS + 2% FBS and Brilliant buffer (BD Bioscience). Antigen probes were prepared individually and combined after multimerization with 5 μM free D-biotin (Avidity) to minimize potential cross-reactivity between probes. For staining, 2–10 × 10^6^ PBMCs per sample were thawed from cryopreservation and prepared in a 96-well round-bottom plate. Cells were first stained with Fc block (BioLegend, 1:200) and Ghost Violet 510 Viability Dye (Tonbo) for 15 min at 4 °C. Cells were then washed and stained for 1 h at 4 °C with 50 μl antigen probe master mix containing the above probes combined immediately before staining. Following this incubation period, cells were washed again and stained with anti-CD27-BUV395, anti-CD3-BUV563, anti-CD38-BUV661, anti-IgD-BV480, anti-CD19-BV750, anti-IgA-FITC, anti-CD21-PE-CF594, anti-IgG-AF700 and anti-CD71-APC-H7 for 30 min at 4 °C. After surface stain, cells were washed and fixed in 1× Stabilizing Fixative (BD Biosciences) overnight at 4 °C.

### Data Analysis

All data were analyzed using Graphpad Prism version 10.1.1 or R version 4.3.2 and visualized using R studio. PyMOL version 2.5.5 was used to visualize ancestral SARS-CoV-2 RBD. Custom R scripts are available upon request.

## Notes

### Author Declarations

IRB of the University of Pennsylvania gave ethical approval for this work.

