## Supplemental Data for "Immunological imprinting shapes the specificity of human antibody responses against SARS-CoV-2 variants"

Supplemental Tables 1-2
Supplemental Figures 1-6

**Supplemental Table 1 Cohort Demographics**

| | BA.5 1st | BA.5 2nd | XBB Infection | XBB.1.5 Vaccination |
|---|---|---|---|---|
| **N** | 7 | 9 | 10 | 15 |
| **Age** | | | | |
| age min | 25 | 25 | 24 | 22 |
| age max | 44 | 59 | 39 | 50 |
| age mean | 31.7 | 39 | 31.3 | 30.2 |
| **Sex** | | | | |
| M | 2 | 3 | 2 | 6 |
| F | 5 | 6 | 8 | 9 |
| **Race** | | | | |
| White | 6 | 6 | 7 | 12 |
| Black | 0 | 1 | 2 | 1 |
| Asian | 1 | 2 | 1 | 2 |
| **Ethnicity** | | | | |
| NHL | 7 | 9 | 9 | 14 |
| HL | 0 | 0 | 1 | 1 |

### Supplementary Table 2: Key Resources Table

| REAGENT or RESOURCE | SOURCE | IDENTIFIER |
|---|---|---|
| Antibodies | | |
| BUV563 anti-CD3 | BD Biosciences | Cat#748569 |
| BV750 anti-CD19 | Biolegend | Cat#302262 |
| BUV805 anti-CD20 | BD Biosciences | Cat#612905 |
| BUV395 anti-CD27 | BD Biosciences | Cat#563815 |
| BUV661 anti-CD38 | BD Biosciences | Cat#612969 |
| APC-H7 anti-CD71 | BD Biosciences | Cat#563671 |
| AF700 anti-CD11c | Biolegend | Cat#337220 |
| FITC anti-IgA | Miltenyi | Cat#130-113-475 |
| BV480 anti-IgD | BD Biosciences | Cat#566138 |
| PE/Dazzle 594 anti-CD21 | Biolegend | Cat#354922 |
| PE-Cy7 anti-IgG | Biolegend | Cat#410722 |
| Chemicals, peptides, and recombinant proteins | | |
| SARS-CoV-2 Biotyinlated Full Length Spike | R&D Systems | Cat#AVI10549-050 |
| HA(ΔTM)(A/Brisbane/02/2018)(H1N1) | Immune Tech | Cat#IT-003-00110ΔTMp |
| HA(ΔTM)(B/Colorado/06/2017) | Immune Tech | Cat#IT-003-B21ΔTMp |
| SARS-CoV-2 Biotyinlated RBD (Ancestral) | Acro Biosystems | Cat#SPD-C82E9-25ug |
| SARS-CoV-2 Biotyinlated RBD (Omicron BA.5) | Acro Biosystems | Cat#SPD-C82Ew-25ug |
| SARS-CoV-2 Biotyinlated RBD (Omicron XBB.1.5) | Acro Biosystems | Cat#SPD-C82Q3-25ug |
| SARS-CoV-2 Biotyinlated N-Terminal Domain | Sino Biological | Cat#40591-V49H-B |
| SARS-CoV-2 Biotyinlated S2 | Acro Biosystems | Cat#S2N-C52E8-25ug |
| SARS-CoV-2 Biotyinlated Nucleocapsid | R&D Systems | Cat#BT10474-050 |
| BV421 Streptavidin | Biolegend | Cat#405226 |
| BV605 Streptavidin | Biolegend | Cat#405229 |
| BV711 Streptavidin | BD Biosciences | Cat#563262 |
| BV786 Streptavidin | BD Biosciences | Cat#563858 |
| BUV615 Streptavidin | BD Biosciences | Cat#613013 |
| BUV737 Streptavidin | BD Biosciences | Cat#612775 |
| PE Streptavidin | Biolegend | Cat#405203 |
| PE-Cy7 Streptavidin | Biolegend | Cat#405206 |
| APC Streptavidin | Biolegend | Cat#405207 |
| Ghost Viability Dye Violet 510 | Tonbo | Cat#13-0870-T100 |
| Human TruStain FcX (Fc Receptor Blocking Solution) | Biolegend | Cat#422302 |

### Antibody Dilutions for Flow Cytometry

| REAGENT or RESOURCE | SOURCE | Dilution Factor | Clone | IDENTIFIER |
|---|---|---|---|---|
| BUV563 anti-CD3 | BD Biosciences | 200 | UCHT1 | Cat#748569 |
| BV750 anti-CD19 | Biolegend | 100 | HIB19 | Cat#302262 |
| BUV395 anti-CD27 | BD Biosciences | 200 | L128 | Cat#563815 |
| BUV661 anti-CD38 | BD Biosciences | 1000 | HIT2 | Cat#612969 |
| APC-H7 anti-CD71 | BD Biosciences | 50 | M-A712 | Cat#563671 |
| FITC anti-IgA | Miltenyi | 400 | IS11-8E10 | Cat#130-113-475 |

| | | | | |
|---|---|---|---|---|
| BV480 anti-IgD | BD Biosciences | 50 | IA6-2 | Cat#566138 |
| PE/Dazzle 594 anti-CD21 | Biolegend | 500 | Bu32 | Cat#354922 |
| AF700 anti-IgG | BD | 200 | G18-145 | Cat#561296 |

S1A

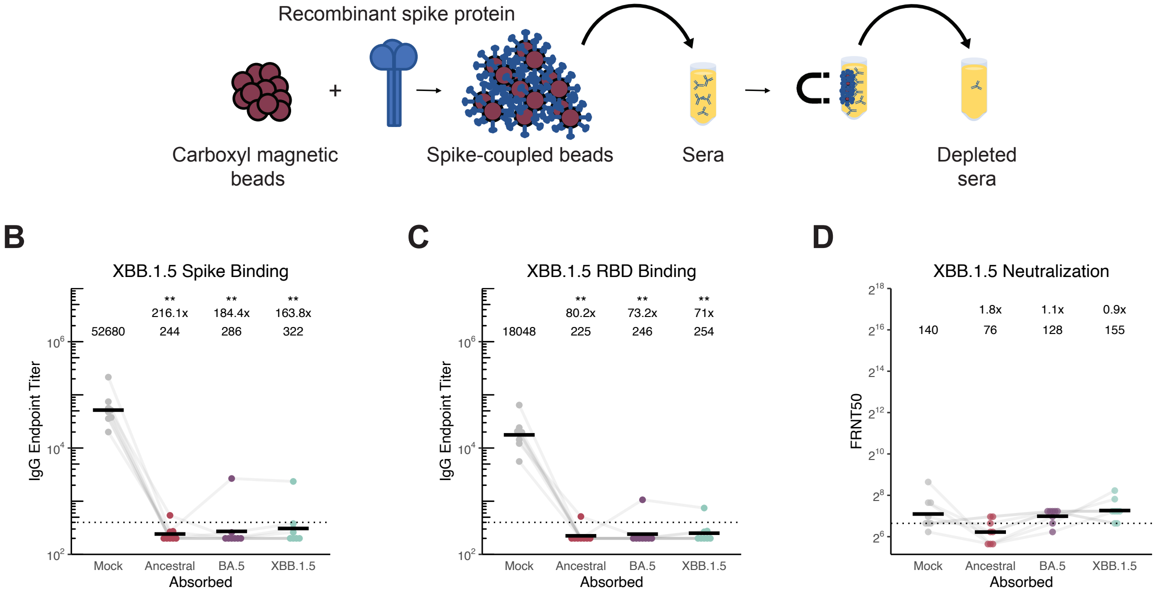
Recombinant spike protein
Carboxyl magnetic beads
Spike-coupled beads
Sera
Depleted sera
B
XBB.1.5 Spike Binding
IgG Endpoint Titer
52680
** 216.1x 244
** 184.4x 286
** 163.8x 322
Mock
Ancestral
BA.5
XBB.1.5
Absorbed
C
XBB.1.5 RBD Binding
IgG Endpoint Titer
18048
** 80.2x 225
** 73.2x 246
** 71x 254
Mock
Ancestral
BA.5
XBB.1.5
Absorbed
D
XBB.1.5 Neutralization
FRNT50
140
1.8x 76
1.1x 128
0.9x 155
Mock
Ancestral
BA.5
XBB.1.5
Absorbed

**Fig S1.** Absorption assay schematic and extended breakthrough cohort absorption analyses. A) Absorption assay workflow. B)-D) Absorbed sera antibodies binding to XBB.1.5 spike (B), RBD (C), and XBB.1.5 pseudovirus neutralization (D). For all, individual points are average of n = 2 technical replicates. Black bars indicate geometric mean. Wilcoxon signed-rank test with benjamini-hochberg correction for multiple testing. All comparisons to timepoint 1 or mock absorption. * $p<0.05$, ** $p<0.01$.

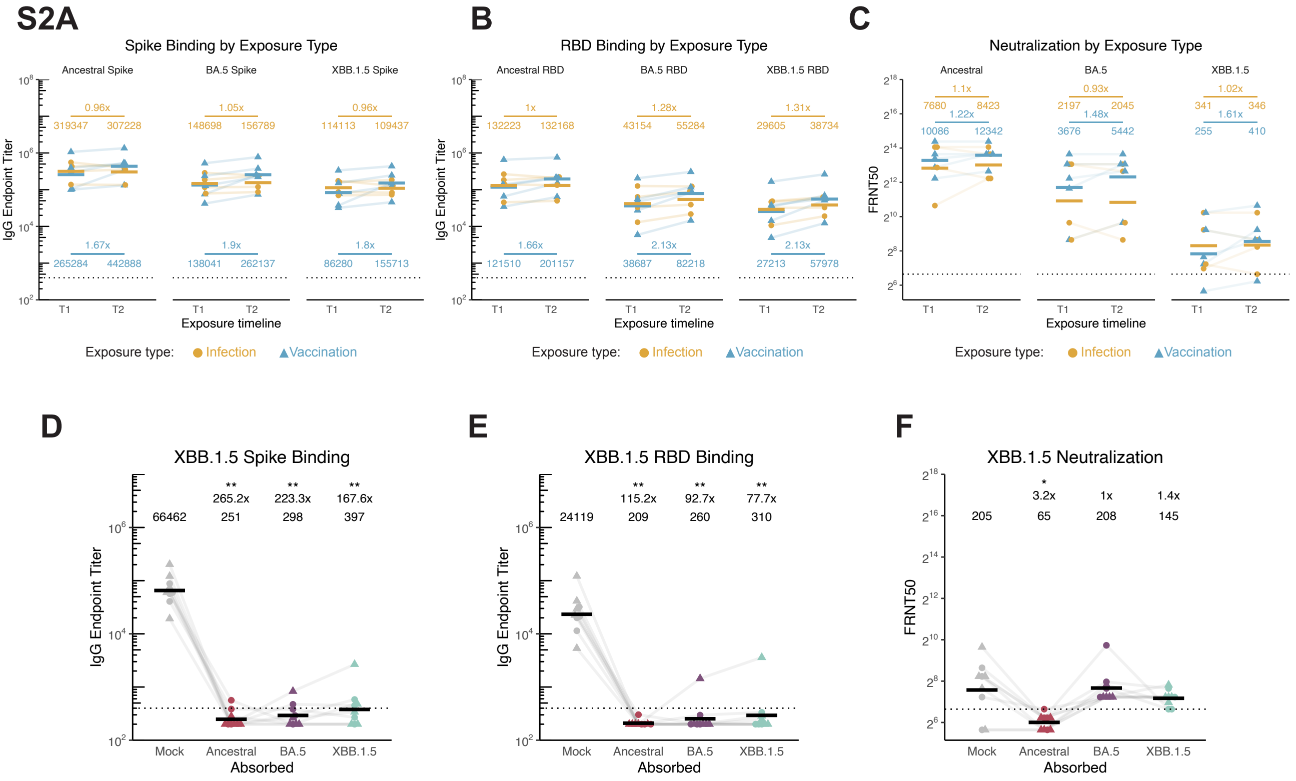
S2A
Spike Binding by Exposure Type
Ancestral Spike
BA.5 Spike
XBB.1.5 Spike
IgG Endpoint Titer
Exposure timeline
B
RBD Binding by Exposure Type
Ancestral RBD
BA.5 RBD
XBB.1.5 RBD
C
Neutralization by Exposure Type
Ancestral
BA.5
XBB.1.5
FRNT50
Exposure type: Infection Vaccination
D
XBB.1.5 Spike Binding
66462
265.2x
251
223.3x
298
167.6x
397
E
XBB.1.5 RBD Binding
24119
115.2x
209
92.7x
260
77.7x
310
F
XBB.1.5 Neutralization
205
3.2x
65
1x
208
1.4x
145
Mock
Ancestral
BA.5
XBB.1.5
Absorbed

**Fig S2.** Extended secondary BA.5 exposure absorption analyses and cohort comparisons. A)-C) Comparison of last BA.5 exposure type (infection or bivalent booster) within the cohort that was exposed twice with BA.5. Spike (A), RBD (B), and pseudovirus neutralization (C) titers compared between infection (gold circle) or bivalent booster (blue triangle) last BA.5 exposures. D)-F) Absorbed sera binding to XBB.1.5 spike (D), RBD (E), and XBB.1.5 pseudovirus neutralization (F). For all, Individual points are average of n = 2 technical replicates. Crossbars indicate geometric mean. Wilcoxon signed-rank test with benjamini-hochberg correction for multiple testing. All comparisons to timepoint 1 or mock absorption. * $p<0.05$, ** $p<0.01$.

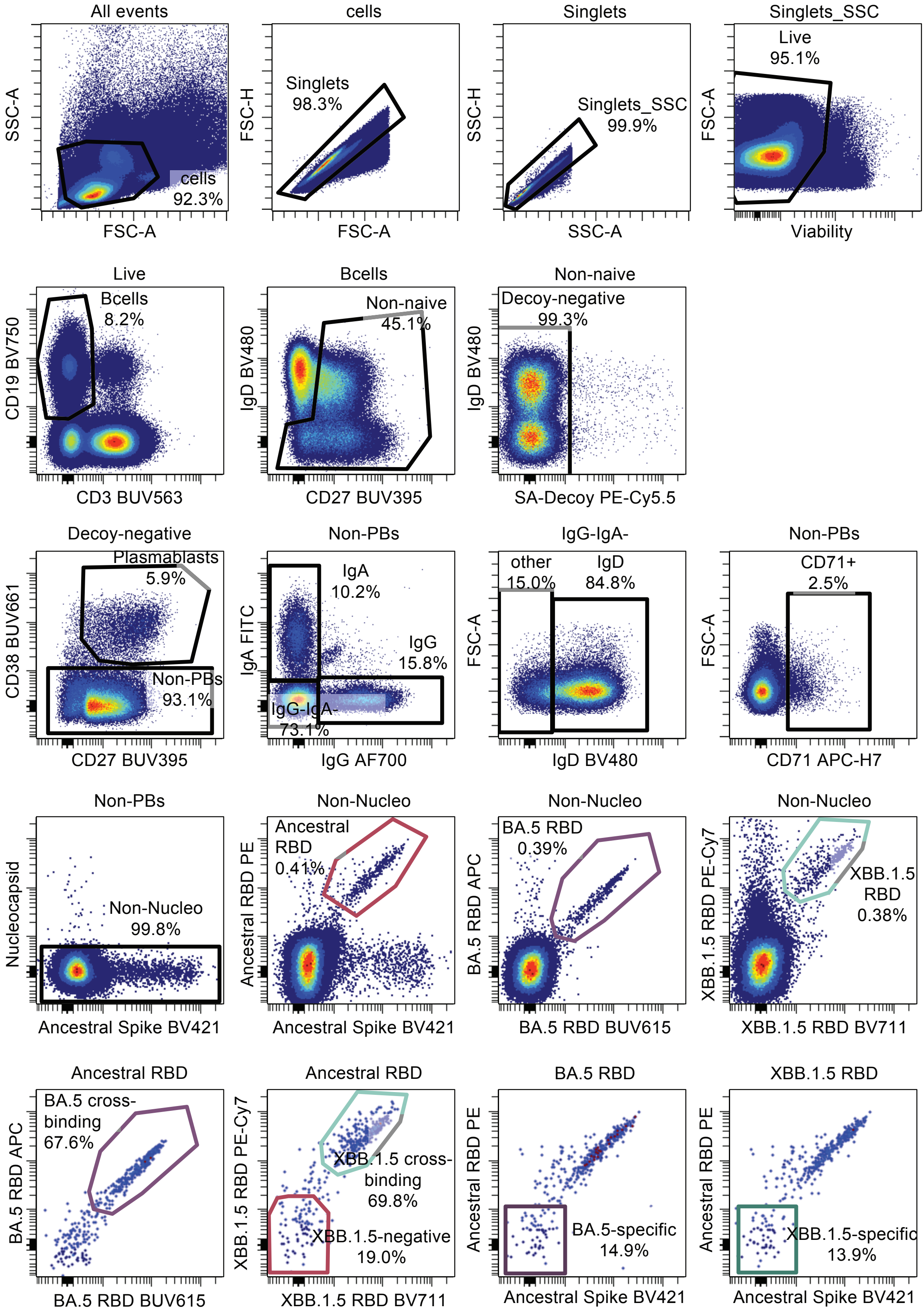
All events
cells
92.3%
SSC-A
FSC-A
cells
Singlets
98.3%
FSC-H
FSC-A
Singlets
Singlets_SSC
99.9%
SSC-H
SSC-A
Singlets_SSC
Live
95.1%
FSC-A
Viability
Live
Bcells
8.2%
CD19 BV750
CD3 BUV563
Bcells
Non-naive
45.1%
IgD BV480
CD27 BUV395
Non-naive
Decoy-negative
99.3%
IgD BV480
SA-Decoy PE-Cy5.5
Decoy-negative
Plasmablasts
5.9%
Non-PBs
93.1%
CD38 BUV661
CD27 BUV395
Non-PBs
IgA
10.2%
IgG
15.8%
IgG-IgA-
73.1%
IgA FITC
IgG AF700
IgG-IgA-
other
15.0%
IgD
84.8%
FSC-A
IgD BV480
Non-PBs
CD71+
2.5%
FSC-A
CD71 APC-H7
Non-PBs
Non-Nucleo
99.8%
Nucleocapsid
Ancestral Spike BV421
Non-Nucleo
Ancestral
RBD
0.41%
Ancestral RBD PE
Ancestral Spike BV421
Non-Nucleo
BA.5 RBD
0.39%
BA.5 RBD APC
BA.5 RBD BUV615
Non-Nucleo
XBB.1.5
RBD
0.38%
XBB.1.5 RBD PE-Cy7
XBB.1.5 RBD BV711
Ancestral RBD
BA.5 cross-
binding
67.6%
BA.5 RBD APC
BA.5 RBD BUV615
Ancestral RBD
XBB.1.5 cross-
binding
69.8%
XBB.1.5-negative
19.0%
XBB.1.5 RBD PE-Cy7
XBB.1.5 RBD BV711
BA.5 RBD
BA.5-specific
14.9%
Ancestral RBD PE
Ancestral Spike BV421
XBB.1.5 RBD
XBB.1.5-specific
13.9%
Ancestral RBD PE
Ancestral Spike BV421

**Fig S3.** Gating strategy for flow cytometric assays using B cell probes. The parent gate is indicated above each plot. All plots are taken from the same representative sample at 15 days post-XBB.1.5 vaccination.

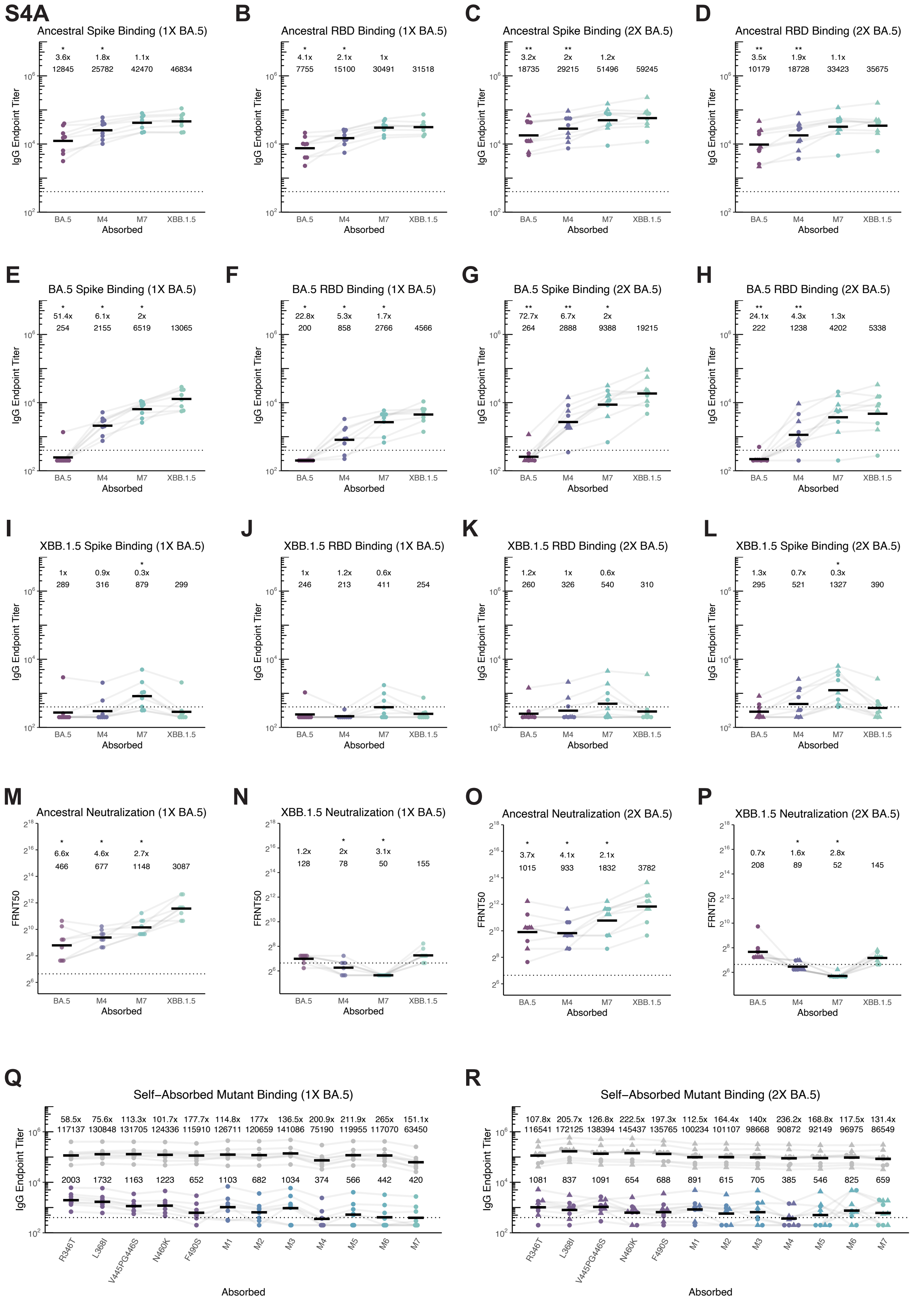
S4A
Ancestral Spike Binding (1X BA.5)
B
Ancestral RBD Binding (1X BA.5)
C
Ancestral Spike Binding (2X BA.5)
D
Ancestral RBD Binding (2X BA.5)
E
BA.5 Spike Binding (1X BA.5)
F
BA.5 RBD Binding (1X BA.5)
G
BA.5 Spike Binding (2X BA.5)
H
BA.5 RBD Binding (2X BA.5)
I
XBB.1.5 Spike Binding (1X BA.5)
J
XBB.1.5 RBD Binding (1X BA.5)
K
XBB.1.5 RBD Binding (2X BA.5)
L
XBB.1.5 Spike Binding (2X BA.5)
M
Ancestral Neutralization (1X BA.5)
N
XBB.1.5 Neutralization (1X BA.5)
O
Ancestral Neutralization (2X BA.5)
P
XBB.1.5 Neutralization (2X BA.5)
Q
Self-Absorbed Mutant Binding (1X BA.5)
R
Self-Absorbed Mutant Binding (2X BA.5)
IgG Endpoint Titer
FRNT50
Absorbed
BA.5
M4
M7
XBB.1.5
R346T
L368I
V445PG446S
N460K
F490S
M1
M2
M3
M5
M6

**Fig S4**. Extended analyses of epitope mapping experiments. A)-P) Binding and neutralization analyses of M4 and M7 absorbed sera using ancestral, BA.5, and XBB.1.5 proteins and pseudoviruses in comparisons to BA.5 and XBB.1.5 absorbed sera. Q),R) Self-absorption controls for mutant spike proteins for sera from 1x (Q) and 2x (R) BA.5 exposed individuals. Self-absorption is represented as the fold reduction in X antigen binding after X antigen absorption (colored points), compared to mock binding (gray points). For all, individual points are average of n = 2 technical replicates. Red/black bars indicate geometric mean. Wilcoxon signed-rank test with benjamini-hochberg correction for multiple testing. A)-P) * $p<0.05$, ** $p<0.01$.

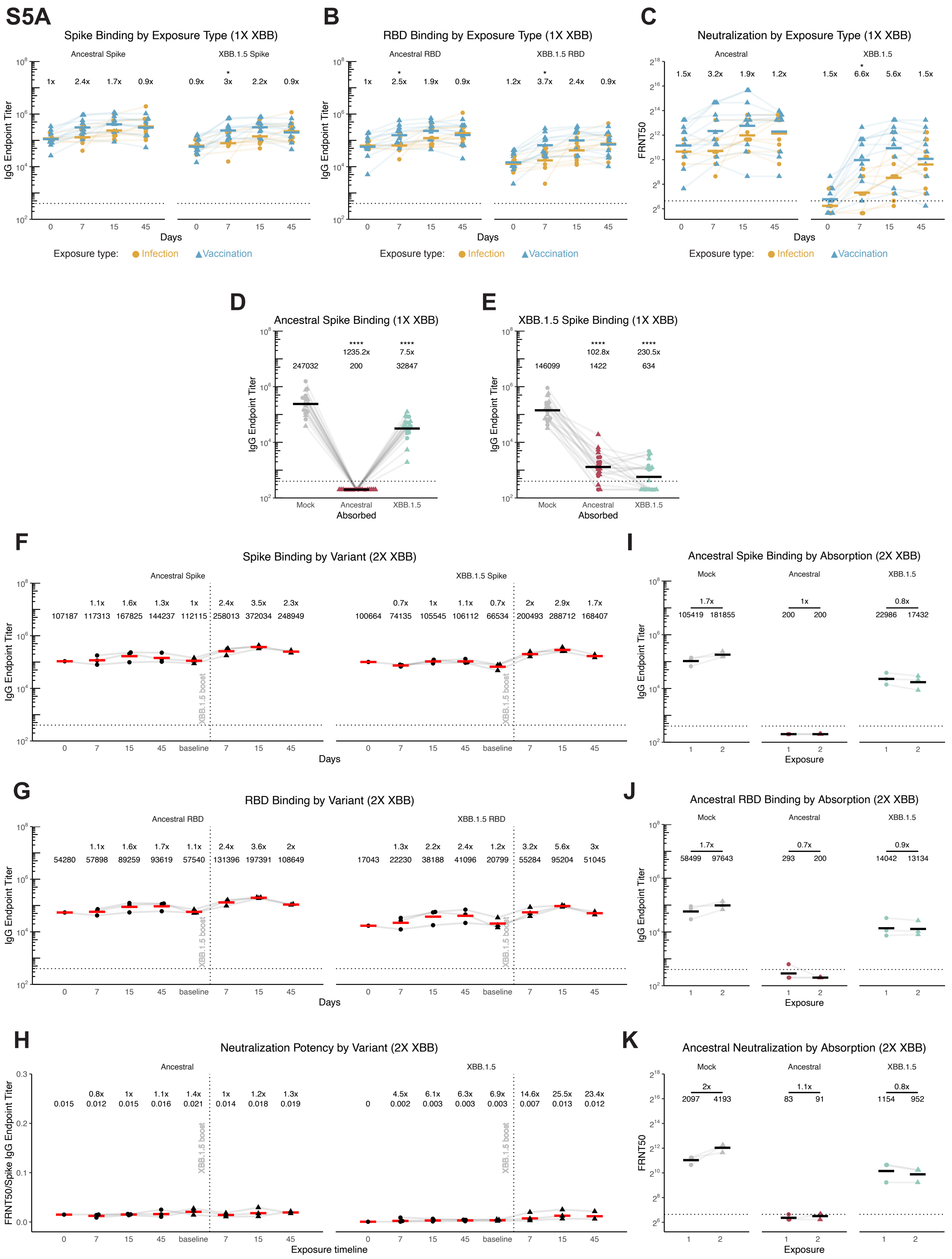
S5A
Spike Binding by Exposure Type (1X XBB)
Ancestral Spike
XBB.1.5 Spike
1x 2.4x 1.7x 0.9x
0.9x 3x 2.2x 0.9x
IgG Endpoint Titer
Days
Exposure type: Infection Vaccination
B
RBD Binding by Exposure Type (1X XBB)
Ancestral RBD
XBB.1.5 RBD
1x 2.5x 1.9x 0.9x
1.2x 3.7x 2.4x 0.9x
C
Neutralization by Exposure Type (1X XBB)
Ancestral
XBB.1.5
1.5x 3.2x 1.9x 1.2x
1.5x 6.6x 5.6x 1.5x
FRNT50
D
Ancestral Spike Binding (1X XBB)
1235.2x 7.5x
247032 200 32847
Mock Ancestral XBB.1.5
Absorbed
E
XBB.1.5 Spike Binding (1X XBB)
102.8x 230.5x
146099 1422 634
F
Spike Binding by Variant (2X XBB)
1.1x 1.6x 1.3x 1x 2.4x 3.5x 2.3x
107187 117313 167825 144237 112115 258013 372034 248949
0.7x 1x 1.1x 0.7x 2x 2.9x 1.7x
100664 74135 105545 106112 66534 200493 288712 168407
XBB.1.5 boost
0 7 15 45 baseline 7 15 45
I
Ancestral Spike Binding by Absorption (2X XBB)
Mock Ancestral XBB.1.5
1.7x 1x 0.8x
105419 181855 200 200 22986 17432
Exposure
G
RBD Binding by Variant (2X XBB)
Ancestral RBD
XBB.1.5 RBD
1.1x 1.6x 1.7x 1.1x 2.4x 3.6x 2x
54280 57898 89259 93619 57540 131396 197391 108649
1.3x 2.2x 2.4x 1.2x 3.2x 5.6x 3x
17043 22230 38188 41096 20799 55284 95204 51045
J
Ancestral RBD Binding by Absorption (2X XBB)
1.7x 0.7x 0.9x
58499 97643 293 200 14042 13134
H
Neutralization Potency by Variant (2X XBB)
0.8x 1x 1.1x 1.4x 1x 1.2x 1.3x
0.015 0.012 0.015 0.016 0.021 0.014 0.018 0.019
4.5x 6.1x 6.3x 6.9x 14.6x 25.5x 23.4x
0 0.002 0.003 0.003 0.003 0.007 0.013 0.012
FRNT50/Spike IgG Endpoint Titer
Exposure timeline
K
Ancestral Neutralization by Absorption (2X XBB)
2x 1.1x 0.8x
2097 4193 83 91 1154 952

**Fig S5.** Extended analyses of XBB cohorts. A)-C) Spike (A) and RBD (B) IgG binding and pseudovirus neutralization (C) comparison between XBB infection (gold circle) and XBB.1.5 vaccination (blue triangle) groups. Ratio at each timepoint is vaccination geometric mean / infection geometric mean. D),E) Ancestral (D) and XBB.1.5 (E) spike IgG binding titers of absorbed sera. F)-H) Spike binding (F), RBD binding (G), and neutralization potency (H) of sera from individuals exposed twice with XBB.1.5. I)-K) Ancestral spike (I), RBD (J), and pseudovirus neutralization (K) titers of sera from individuals exposed twice with XBB.1.5.. For all, individual points are average of n = 2 technical replicates. Red/black bars indicate geometric mean. Wilcoxon rank-sum test with benjamini-hochberg correction for multiple testing. All comparisons to timepoint 1 or mock absorption. * $p<0.05$, **** $p<0.0001$.

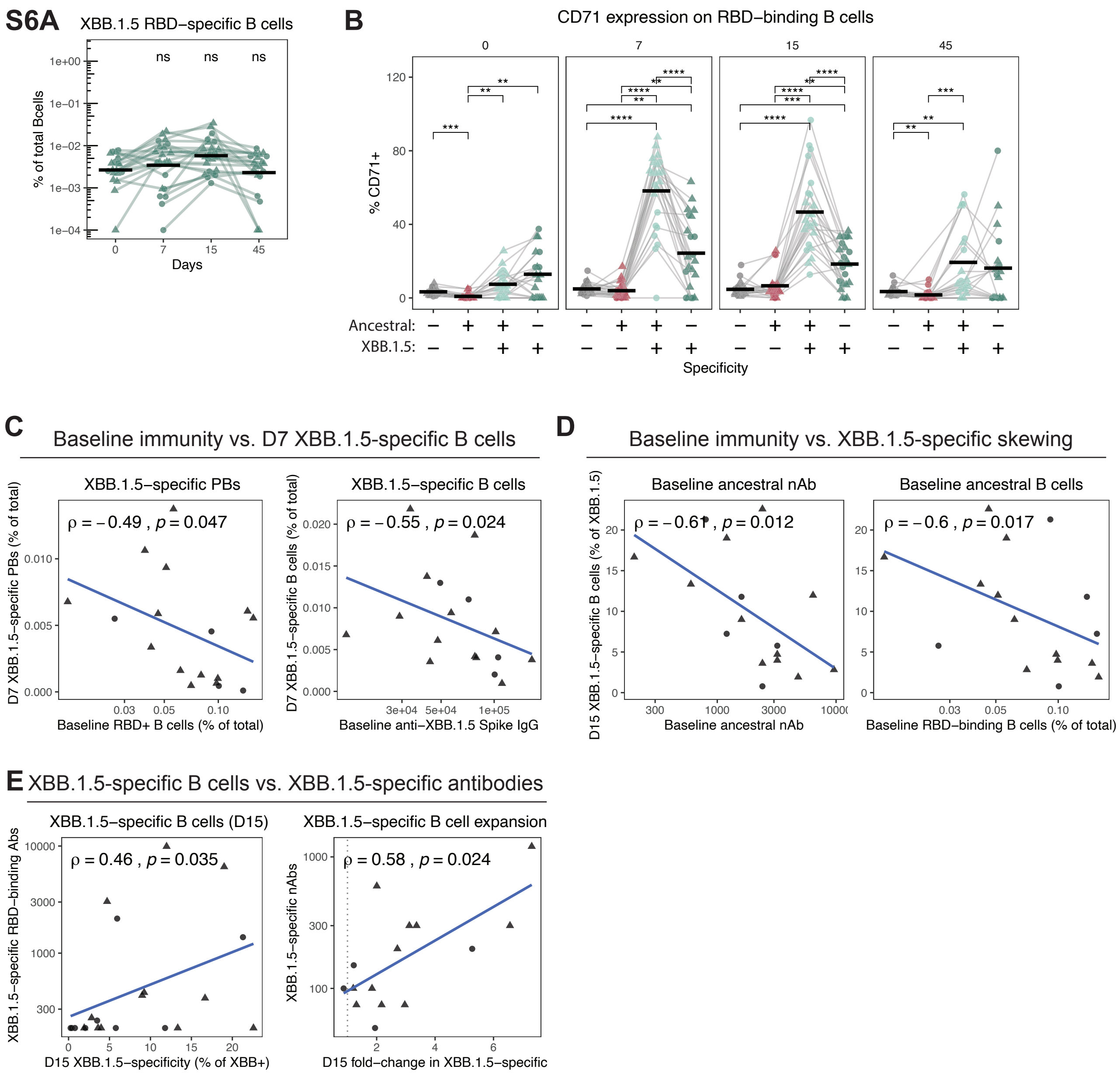
S6A
XBB.1.5 RBD−specific B cells
% of total Bcells
Days
ns
B
CD71 expression on RBD−binding B cells
% CD71+
Ancestral:
XBB.1.5:
Specificity
C
Baseline immunity vs. D7 XBB.1.5-specific B cells
XBB.1.5−specific PBs
ρ = −0.49 , p = 0.047
D7 XBB.1.5−specific PBs (% of total)
Baseline RBD+ B cells (% of total)
XBB.1.5−specific B cells
ρ = −0.55 , p = 0.024
D7 XBB.1.5−specific B cells (% of total)
Baseline anti−XBB.1.5 Spike IgG
D
Baseline immunity vs. XBB.1.5-specific skewing
Baseline ancestral nAb
ρ = −0.61 , p = 0.012
D15 XBB.1.5−specific B cells (% of XBB.1.5)
Baseline ancestral nAb
Baseline ancestral B cells
ρ = −0.6 , p = 0.017
Baseline RBD−binding B cells (% of total)
E
XBB.1.5-specific B cells vs. XBB.1.5-specific antibodies
XBB.1.5−specific B cells (D15)
ρ = 0.46 , p = 0.035
XBB.1.5−specific RBD−binding Abs
D15 XBB.1.5−specificity (% of XBB+)
XBB.1.5−specific B cell expansion
ρ = 0.58 , p = 0.024
XBB.1.5−specific nAbs
D15 fold−change in XBB.1.5−specific

**Fig S6.** Extended flow cytometric analyses of XBB cohorts. A) The percent of total B cells that bind XBB.1.5 RBD but not ancestral RBD before and after exposure to XBB. B) Summary data of percent of the indicated RBD-binding B cell populations that express CD71 at 0, 7, 15, and 45 days post-XBB exposure. C-D) Correlations within B cell and antibody responses. C) Correlation of day 0 percent of total B cells that bind ancestral RBD and day 7 percent of total B cells that are CD27+CD38+ plasmablasts that bind XBB.1.5 RBD but not ancestral RBD (XBB.1.5-specific, left), and day 0 XBB.1.5 Spike binding antibodies by ELISA and day 7 percent of total B cells that bind XBB.1.5 RBD but not ancestral RBD (XBB.1.5-specific, right). D) Correlation of day 15 percent of XBB.1.5 RBD-binding B cells that do not bind ancestral RBD with day 0 ancestral neutralizing antibody titers (left) and day 0 percent of total B cells that bind ancestral RBD (right). Points represent individual subjects and thin lines indicate individual subjects sampled longitudinally. Horizontal bars represent means. Statistics were calculated using two-sided Wilcoxon test with Benjamini-Hochberg correction for multiple comparisons. Statistics without brackets are in comparison to day 0. Correlation statistics were calculated using Spearman rank correlation and are shown with Pearson trend lines for visualization.
